# COVID-19 vaccination, risk-compensatory behaviours, and contacts in the UK

**DOI:** 10.1101/2021.11.15.21266255

**Authors:** John Buckell, Joel Jones, Philippa C Matthews, Sir Ian Diamond, Emma Rourke, Ruth Studley, Duncan Cook, A Sarah Walker, Koen B Pouwels, the COVID-19 Infection Survey Team

## Abstract

The physiological effects of vaccination against SARS-CoV-2 (COVID-19) are well documented, yet the behavioural effects are largely unknown. Risk compensation suggests that gains in personal safety, as a result of vaccination, are offset by increases in risky behaviour, such as socialising, commuting and working outside the home. This is potentially problematic because transmission of SARS-CoV-2 is driven by contacts, which could be amplified by vaccine-related risk compensation behaviours. Here, we show that behaviours were overall unrelated to personal vaccination, but - adjusting for variation in mitigation policies - were responsive to the level of vaccination in the wider population: individuals in the UK were risk compensating when rates of vaccination were rising. This effect was observed across four nations of the UK, each of which varied policies autonomously.

## Introduction

The United Kingdom was the first country to begin a national COVID-19 vaccination programme on 8 December 2020, and as of 15^th^ October 2021, 86% of the adult population (aged 18 and over) had received a first dose^1^.

In the UK, the most commonly used vaccines are the BNT162b2 messenger RNA (mRNA) vaccine (Pfizer-BioNTech), the mRNA-1273 (Moderna) vaccine, and the Oxford-AstraZeneca adenovirus vector vaccine, ChAdOx1 nCOV-19 (denoted here ChAdOx1). These vaccines are effective in preventing (symptomatic) SARS-CoV-2 infection and hospitalisation and deaths^2-10^. While the protective effect against infection and onward transmission wanes after vaccination and is slightly reduced with the Delta variant compared to the Alpha variant^11-12^, protection against severe outcomes remains high in most studies^8^.

Whether SARS-CoV-2 vaccination induces a behavioural, as well as physiological, response in individuals is not known. Individuals who have been immunised may be less fearful about contracting SARS-CoV-2, getting severely ill from infection, or spreading the virus when getting infected. In turn, they may begin to interact with others more often or less cautiously. This type of behaviour is known as risk compensation or the Peltzman effect^13^, and has been studied in various health settings with mixed evidence on its presence^14-16^. In the context of SARS-CoV-2, there is scepticism regarding risk compensation for face coverings^17^, but concern that risk compensation may pervade post-vaccination behaviour^18,19^.

Risk compensation could present significant short- and medium-term public health risks, particularly if individuals – or their unvaccinated household members – change their behaviour before being fully protected by vaccination or when protection from vaccination is incomplete or temporary. Misconceptions about the extent of protection after COVID-19 vaccination might be common, as exemplified by the Dutch health minister recently advocating that one could go partying one day after getting a single dose of the Ad26.COV2.S (Janssen) vaccine^20^. Developing an understanding of the extent to which post-vaccination behaviours are risk-compensatory is important for public health messaging and policy development.

We used the Office for National Statistics (ONS) COVID-19 Infection Survey (CIS) – a large community-based survey of individuals aged 2 years and older living in randomly selected households across the United Kingdom – to evaluate behaviours of individuals in response to vaccine uptake by themselves, their vulnerable household members, and in their specific geographical area (CIS geographical regions, which is 133 geographical areas in England, Northern Ireland, Scotland, and Wales). We investigate risk-compensatory behaviours in 10 reported outcomes, including physical and socially-distanced contacts (each divided into three categories according to the age of the contact), time that others spend in individuals’ homes, individuals’ time in other people’s houses, location of work (home, outside the home, combination), and private vs public transport for commuting.

## Results

### Behaviour changes among vaccinated individuals aged 18-64 years

This analysis included observations between 1^st^ October 2020 and 15^th^ September 2021 representing 1,839,911 visits of 161,309 individuals aged 18 to 64 years from 102,260 households who did not report working in a patient-facing healthcare role. 18 to 64 year-olds were selected as the vaccination program had offered vaccines to individuals of these ages at the time of analysis, which was not true of younger ages. Individuals were only included if there was evidence that they were vaccinated at any point during the survey to avoid that those not willing to get vaccinated, who are expected to have different behaviours than those that are willing to get vaccinated. We excluded over-65s (because patterns of behaviour were different) and those reporting working in patient-facing healthcare (see appendix 2). Characteristics of included individuals are summarized in Table 1.

**Table 1:** Descriptive statistics of the characteristics of individuals included in analyses at the first visit on or after 1 October 2020.

|  | Own vaccination 18 to 64y<br>N = 161,309 |  |  |  | Household vaccination 18 to 64y<br>N = 7,626 |  |  |  | Population vaccination 2+ years<br>N = 501,679 |  |  |  |
| --- | --- | --- | --- | --- | --- | --- | --- | --- | --- | --- | --- | --- |
|  | Mean | s.d. | Min | Max | Mean | s.d. | Min | Max | Mean | s.d. | Min | Max |
| Age | 45.57 | 12.44 | 18.00 | 64.00 | 46.28 | 14.21 | 18.00 | 64.00 | 47.12 | 21.81 | 2.00 | 95.00 |
| Female | 0.53 | 0.50 | 0.00 | 1.00 | 0.53 | 0.50 | 0.00 | 1.00 | 0.53 | 0.50 | 0.00 | 1.00 |
| Black | 0.01 | 0.09 | 0.00 | 1.00 | 0.01 | 0.08 | 0.00 | 1.00 | 0.01 | 0.10 | 0.00 | 1.00 |
| Asian | 0.05 | 0.21 | 0.00 | 1.00 | 0.05 | 0.21 | 0.00 | 1.00 | 0.04 | 0.20 | 0.00 | 1.00 |
| Mixed | 0.01 | 0.12 | 0.00 | 1.00 | 0.02 | 0.13 | 0.00 | 1.00 | 0.02 | 0.13 | 0.00 | 1.00 |
| Other | 0.01 | 0.10 | 0.00 | 1.00 | 0.01 | 0.09 | 0.00 | 1.00 | 0.01 | 0.10 | 0.00 | 1.00 |
| Country IMD Rank | 58.14 | 27.06 | 0.00 | 100.00 | 58.98 | 26.23 | 0.02 | 100.00 | 58.33 | 27.00 | 0.00 | 100.00 |
| North East | 0.04 | 0.20 | 0.00 | 1.00 | 0.04 | 0.20 | 0.00 | 1.00 | 0.04 | 0.19 | 0.00 | 1.00 |
| North West | 0.12 | 0.33 | 0.00 | 1.00 | 0.12 | 0.33 | 0.00 | 1.00 | 0.12 | 0.32 | 0.00 | 1.00 |
| Yorkshire & The Humber | 0.09 | 0.28 | 0.00 | 1.00 | 0.08 | 0.28 | 0.00 | 1.00 | 0.08 | 0.28 | 0.00 | 1.00 |
| East Midlands | 0.07 | 0.25 | 0.00 | 1.00 | 0.07 | 0.25 | 0.00 | 1.00 | 0.07 | 0.25 | 0.00 | 1.00 |
| West Midlands | 0.08 | 0.27 | 0.00 | 1.00 | 0.09 | 0.28 | 0.00 | 1.00 | 0.08 | 0.27 | 0.00 | 1.00 |
| East | 0.11 | 0.31 | 0.00 | 1.00 | 0.11 | 0.32 | 0.00 | 1.00 | 0.10 | 0.30 | 0.00 | 1.00 |
| London | 0.20 | 0.40 | 0.00 | 1.00 | 0.17 | 0.38 | 0.00 | 1.00 | 0.18 | 0.39 | 0.00 | 1.00 |
| South East | 0.13 | 0.34 | 0.00 | 1.00 | 0.14 | 0.34 | 0.00 | 1.00 | 0.13 | 0.34 | 0.00 | 1.00 |
| South West | 0.08 | 0.26 | 0.00 | 1.00 | 0.09 | 0.28 | 0.00 | 1.00 | 0.08 | 0.28 | 0.00 | 1.00 |
| N Ireland | 0.02 | 0.14 | 0.00 | 1.00 | 0.02 | 0.14 | 0.00 | 1.00 | 0.02 | 0.12 | 0.00 | 1.00 |
| Scotland | 0.04 | 0.19 | 0.00 | 1.00 | 0.04 | 0.20 | 0.00 | 1.00 | 0.08 | 0.28 | 0.00 | 1.00 |
| Wales | 0.03 | 0.17 | 0.00 | 1.00 | 0.03 | 0.17 | 0.00 | 1.00 | 0.02 | 0.15 | 0.00 | 1.00 |
| Household size 2 | 0.40 | 0.49 | 0.00 | 1.00 | 0.55 | 0.50 | 0.00 | 1.00 | 0.41 | 0.49 | 0.00 | 1.00 |
| Household size 3 | 0.20 | 0.40 | 0.00 | 1.00 | 0.26 | 0.44 | 0.00 | 1.00 | 0.17 | 0.37 | 0.00 | 1.00 |
| Household size 4 | 0.18 | 0.39 | 0.00 | 1.00 | 0.14 | 0.34 | 0.00 | 1.00 | 0.17 | 0.38 | 0.00 | 1.00 |
| Household size 5 | 0.07 | 0.25 | 0.00 | 1.00 | 0.06 | 0.23 | 0.00 | 1.00 | 0.08 | 0.28 | 0.00 | 1.00 |
| Multi-generation | 0.06 | 0.24 | 0.00 | 1.00 | 0.05 | 0.22 | 0.00 | 1.00 | 0.07 | 0.25 | 0.00 | 1.00 |
| Urban/rural 2 | 0.42 | 0.49 | 0.00 | 1.00 | 0.42 | 0.49 | 0.00 | 1.00 | 0.42 | 0.49 | 0.00 | 1.00 |
| Urban/rural 3 | 0.10 | 0.30 | 0.00 | 1.00 | 0.11 | 0.31 | 0.00 | 1.00 | 0.10 | 0.31 | 0.00 | 1.00 |
| Urban/rural 4 | 0.10 | 0.29 | 0.00 | 1.00 | 0.12 | 0.33 | 0.00 | 1.00 | 0.11 | 0.31 | 0.00 | 1.00 |

IMD – index of multiple deprivation.

The mean outcome values are presented in Table 2 and aggregated into binary variables of “zero [contacts]” or “any [contacts]” (modelling used all outcome categories). On average, individuals were most likely to report having at least one socially-distanced contact with someone aged 18 to 69 years outside their household in the past 7 days (75.5% of visits). Among the risk behaviours considered, on average, individuals were least likely to report having used public transport for a commute (for work or study purposes) in the past 7 days (9.5% of visits). Individuals were slightly more likely to report that others visited their home in the past 7 days (40.8%) than they themselves visiting others’ homes in the past 7 days (36.4%). Individuals were more likely to report working at home than working outside of home over the past 7 days (46.6% reporting only working at home).

**Table 2:** Summaries of self-reported behaviours across all outcomes for in different analyses.

| Outcome | Behaviour captured by binary variable | % of Any |  |  |
| --- | --- | --- | --- | --- |
|  |  | Own vaccination<br>18-64y | Household vaccination<br>18-64y | Population vaccination<br>2+ years |
| Physical contacts with under 18s | Any past 7-day contacts outside the household | 23.5% | 22.4% | 29.7% |
| Physical contacts with 18 to 69 years | Any past 7-day contacts outside the household | 38.1% | 32.9% | 39.7% |
| Physical contacts with over 70s | Any past 7-day contacts outside the household | 17.6% | 14.9% | 18.3% |
| Socially-distanced contacts with under 18s | Any past 7-day contacts outside the household | 34.2% | 30.3% | 36.2% |
| Socially-distanced contacts with 18 to 69 years | Any past 7-day contacts outside the household | 75.5% | 72.1% | 72.2% |
| Socially-distanced contacts with over 70s | Any past 7-day contacts outside the household | 34.2% | 32.6% | 34.4% |
| Visits to others' homes | Any past 7-day home visits | 36.4% | 35.7% | 34.9% |
| Others' visits to own home | Any past 7-day home visits | 40.8% | 40.7% | 40.5% |
| Working/studying outside home | Any past 7-day working outside home | 46.6% | 46.2% | 43.2% |
| Taking public transport to work/place of education | Any past 7-day public transport | 9.5% | 8.7% | 11.0% |

The numerator is the number of observations for which “any” was reported and the denominator is the total number of observations in that analysis. For own vaccination, 23.5% of observations were reported as any past 7-day, outside of the household, physical contacts with under 18 year-olds. All contact variables relate to individuals outside the household. Denominators for work variables restricted to those reporting working or in education. Tables showing how these percentages were derived are provided in appendices 8 to 10.

We calculated the variation in 10 behavioural outcomes as a function of the time to individuals’ own first vaccination (Fig 1), adjusting for multiple confounders, including household composition, individual characteristics, sociodemographic variables, and background prevalence using ordinal generalised additive models. Overall, reported socialising increased over time, with no observed change associated with vaccination.

**Figure 1:**
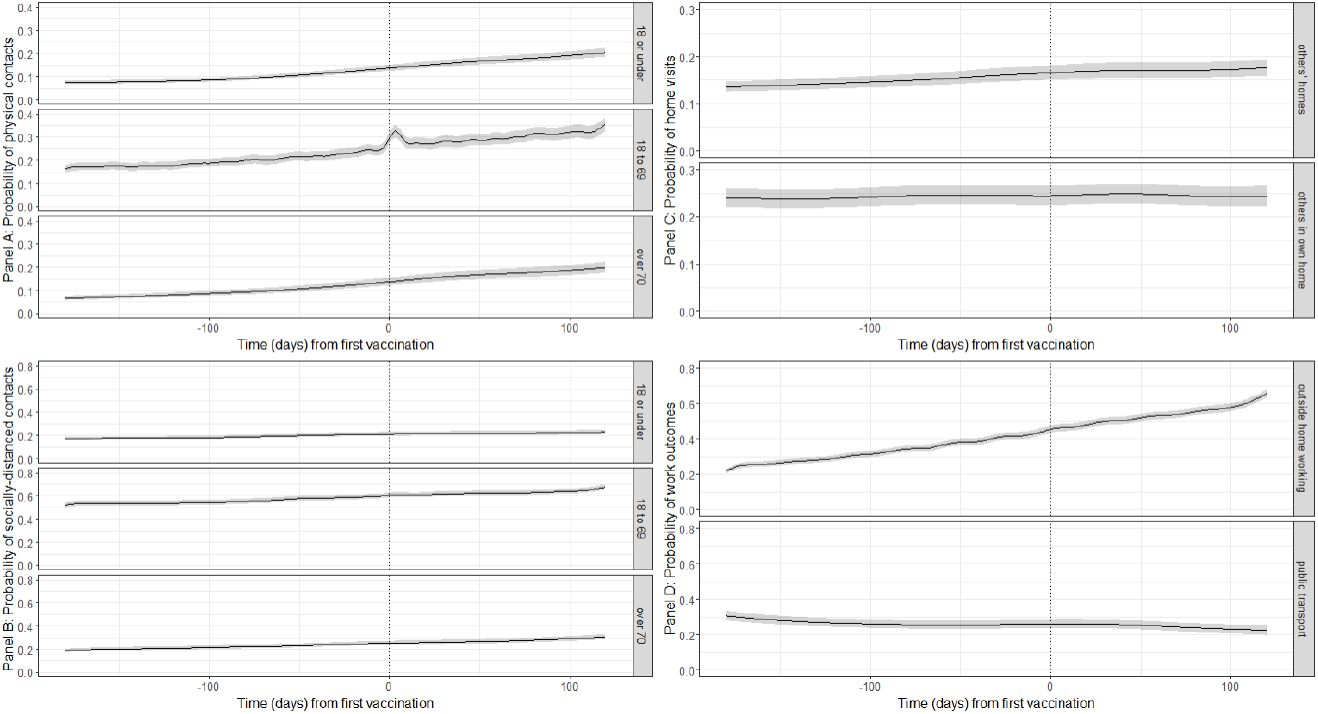
Probabilities of behavioural outcomes for individuals aged 18-64y by time from first vaccination, first dose. Top left (panel A): past 7-day reported physical, outside of household contacts; bottom left (Panel B): past 7-day reported socially-distanced, outside of household contacts; top right (Panel C): past 7-day reported home visits; bottom right (Panel D): past 7-day reported work outcomes for those that are working or in education. Dotted line shows day of own first vaccination. “18 or under”, “18 to 69” and “over 70” denote the ages of the people with whom individuals in the sample had contact.

The probability of any physical contacts increased with time since first vaccination for outside of the household contacts with under-18s, 18 to 69 year-olds, and over-70s (Fig 1A). For physical contacts with either under-18s or over-70s, no difference in the pre- and post-vaccination gradients were observed; that is, there was no evidence of a behavioural response to being vaccinated. To illustrate, for physical contacts with under 18 year-olds, after adjusting for confounders including calendar time, the pre-vaccination gradient for 50 to 14 days prior to vaccination was 0.00048 [95%CI: 0.00040-0.0057] meaning that for each 100 days, the probability of any physical contacts with under 18 year-olds would increase by 0.048. The post-vaccination gradient for the period 1 to 13 days after vaccination was 0.00047 [95% CI 0.00032-0.00058; difference of −0.00004, 95% CI −0.00019 to 0.00012 compared to 50-14 days before]. Thus, the gradients in the two periods appear to be almost identical. For physical contacts with 18 to 69 year-olds, the probability temporarily increased in the immediate period after the first vaccination (Fig 1A & Table 3). However, this increase was only transient, with no evidence of difference in gradient before versus 14 to 50 days after vaccination (Fig 1A & Table 3).

**Table 3:** Gradients and differences in gradients of outcomes over time between pre- and post-vaccination periods, individuals’ own vaccination (18-64y).

| Outcome | Period compared | Gradient (probability) |  | Difference in gradients: percentiles of simulated distribution |  |  |
| --- | --- | --- | --- | --- | --- | --- |
|  |  | Pre vaccination | Post vaccination | 2.5 | 50 | 97.5 |
| Physical contacts with under 18s | -50 to -14 days versus 1 to 13 days | 0.00048 | 0.00047 | -0.00019 | -0.00004 | 0.00012 |
| Physical contacts with 18 to 69 years | -50 to -14 days versus 1 to 6 days | 0.00042 | -0.00025 | -0.00195 | -0.00067 | 0.00059 |
| Physical contacts with 18 to 69 years | -50 to -14 days versus 7 to 13 days | 0.00042 | -0.00367 | -0.00475 | -0.00367 | -0.00264 |
| Physical contacts with over 70s | -50 to -14 days versus 1 to 13 days | 0.00041 | 0.00053 | -0.00003 | 0.00012 | 0.00026 |
| Socially-distanced contacts with under 18s | -50 to -14 days versus 1 to 13 days | 0.00029 | 0.00014 | -0.00037 | -0.00015 | 0.00007 |
| Socially-distanced contacts with 18 to 69 years | -50 to -14 days versus 1 to 13 days | 0.00038 | -0.00020 | -0.00100 | -0.00057 | -0.00014 |
| Socially-distanced contacts with over 70s | -50 to -14 days versus 1 to 13 days | 0.00042 | 0.00029 | -0.00030 | -0.00013 | 0.00005 |
| Visits to others' homes | -50 to -14 days versus 1 to 13 days | 0.00028 | 0.00016 | -0.00021 | -0.00012 | 0.00000 |
| Others' visits to own home | -50 to -14 days versus 1 to 13 days | 0.00003 | 0.00016 | -0.00001 | 0.00013 | 0.00027 |
| Working/studying outside home | -50 to -14 days versus 1 to 13 days | 0.00093 | 0.00063 | -0.00018 | 0.00031 | 0.00079 |
| Taking public transport to work/place of education | -50 to -14 days versus 1 to 13 days | -0.00012 | -0.00012 | -0.00022 | -0.00001 | 0.00023 |
| Physical contacts with under 18s | -50 to -14 days versus 14 to 50 days | 0.00048 | 0.00047 | -0.00012 | -0.00001 | 0.00009 |
| Physical contacts with 18 to 69 years | -50 to -14 days versus 14 to 50 days | 0.00042 | 0.00018 | -0.00048 | -0.00024 | 0.00000 |
| Physical contacts with over 70s | -50 to -14 days versus 14 to 50 days | 0.00041 | 0.00042 | -0.00009 | 0.00000 | 0.00001 |
| Socially-distanced contacts with under 18s | -50 to -14 days versus 14 to 50 days | 0.00029 | 0.00024 | -0.00018 | -0.00004 | 0.00009 |
| Socially-distanced contacts with 18 to 69 years | -50 to -14 days versus 14 to 50 days | 0.00038 | 0.00045 | -0.00013 | 0.00007 | 0.00027 |
| Socially-distanced contacts with over 70s | -50 to -14 days versus 14 to 50 days | 0.00042 | 0.00022 | -0.00031 | -0.00019 | -0.00008 |
| Visits to others' homes | -50 to -14 days versus 14 to 50 days | 0.00028 | 0.00007 | -0.00028 | -0.00021 | -0.00013 |
| Others' visits to own home | -50 to -14 days versus 14 to 50 days | 0.00003 | 0.00013 | -0.00001 | 0.00010 | 0.00021 |
| Working/studying outside home | -50 to -14 days versus 14 to 50 days | 0.00093 | 0.00113 | -0.00042 | -0.00019 | 0.00003 |
| Taking public transport to work/place of education | -50 to -14 days versus 14 to 50 days | -0.00012 | -0.00014 | -0.00022 | -0.00002 | 0.000188 |

Percentiles of the distribution of 10,000 simulations from the Bayesian posterior distributions of the fitted GAM models for each outcome. Pre-vaccination was 50 to 14 days prior to first vaccination. Post-vaccination periods were 1 to 13 days (upper table) and 14 to 50 days (lower table). For 18 to 69 year-old physical contacts, the pre-period was divided into two parts to reflect the peak in probability around the time of vaccination. An example for interpretation: For physical contacts with under 18 year-olds, the pre-vaccination gradient for 50 to 14 days prior to vaccination was 0.0004 meaning that for each 100 days, the probability of any physical contacts with under 18 year-olds would increase by 0.04.

There were no differences between the gradients of the pre- and post-vaccination periods in terms of reporting any socially-distanced contacts with others outside the household (Fig 1B & Table 3), the probability of reporting any visits to others’ home or others’ visits to own home (Fig 1C & Table 3), or the probability of reporting working from home and using public transport for commuting (Fig 1D & Table 3). This was, broadly, the case across the ten behaviours shown in Fig 1 for the comparison of post-vaccination periods with pre-vaccination. Thus, there was no evidence of a behavioural response to being vaccinated on these outcomes.

### Influence of vaccination of vulnerable household members on behaviour of unvaccinated 18-64 year olds

This analysis included observations between 1^st^ October 2020 and 15^th^ September 2021 representing 53,846 visits of 7,626 unvaccinated individuals aged 18 to 64 years old without long-term health conditions and who did not report working in patient-facing healthcare roles from 6,958 households who had at least one vulnerable household member (defined as aged 65 and over or had a long-term health condition, in keeping with vaccine allocation groups of the UK government^21^ (UK government, 2021c)). Characteristics of included individuals are summarized in Table 1. Unvaccinated individuals were included only up until they had themselves been vaccinated, i.e. examining how individuals living with vulnerable household members reacted to the vulnerable household members being vaccinated, before they themselves were vaccinated. We calculated the variation in 10 behavioural outcomes of these individuals, adjusting for multiple confounders including household composition, individual characteristics, sociodemographic variables; and background prevalence (Fig 2). There is some evidence to suggest that physical contacts with under-18s increased following the first vaccination of the last vulnerable household member (Fig 2A). Otherwise, no meaningful difference in the trend was observed between the probability of physical contacts and time to the last vulnerable person in the household having their first vaccination. We found no evidence that trends in other reported behaviours changed upon vaccination of the last vulnerable household member in the household (Fig 2B-D).

**Figure 2:**
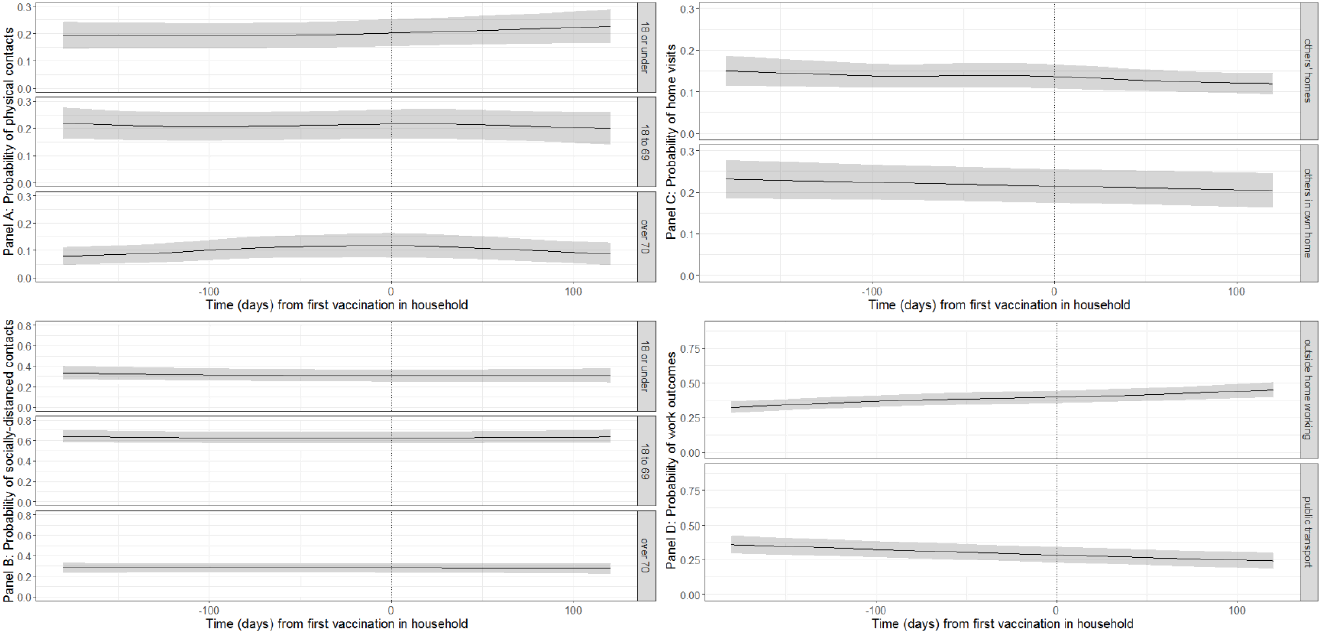
Probabilities of behavioural outcomes for unvaccinated individuals aged 18-64y by time to vaccination of the first vulnerable person in the household, first dose. Top left (panel A): past 7-day reported physical, outside of household contacts; bottom left (Panel B): past 7-day reported socially-distanced, outside of household contacts; top right (Panel C): past 7-day reported home visits; bottom right (Panel D): past 7-day reported work outcomes for those that are working or in education. Dotted line shows day of own first vaccination. “18 or under”, “18 to 69” and “over 70” denote the ages of the people with whom individuals in the sample had contact.

### Influence of population-level vaccine uptake

This analysis included observations between 1^st^ October 2020 and 15^th^ September 2021 representing 4,508,755 visits of 501,679 individuals aged 2 to 95 years old from 238,641 households. Characteristics of included individuals are summarized in Table 1.

Population rates of first vaccination (taken from administrative sources) increased over time during 2021 across regions in England and in Northern Ireland, Scotland, and Wales (Fig 3). Vaccination rates increased earlier in the English regions compared to the other countries in the UK, but from April until October 2021, the percentages were highest in Wales and Scotland, with vaccination rates in London appearing to lag behind other parts of the UK. While there are considerable uncertainties about the actual number of individuals eligible in each area, this is the information that is provided by the Government and the source individuals may act upon if they are responsive to population-level vaccine uptake information.

**Figure 3:**
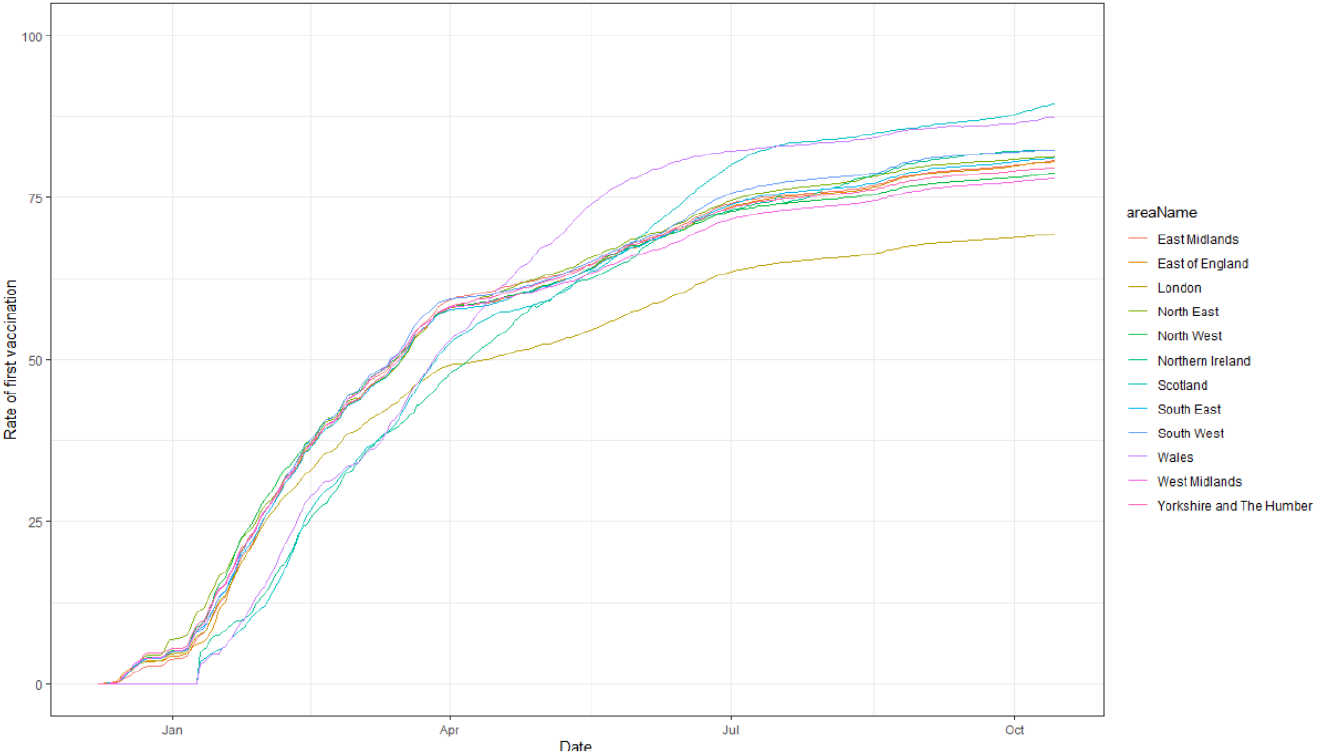
Percentage population SARS-CoV-2 vaccine uptake (first dose) over time during 2021 in all four countries (split by region in England). Source: UK government official statistics, 2021a.

We calculated the variation in 10 behavioural outcomes as a function of the percentage of people who had had a first vaccination in the local population (defined as CIS geographical regions), using all individuals in the survey (Fig 4), adjusting for multiple confounders including household composition, individual characteristics, sociodemographic variables, and background prevalence.

**Figure 4:**
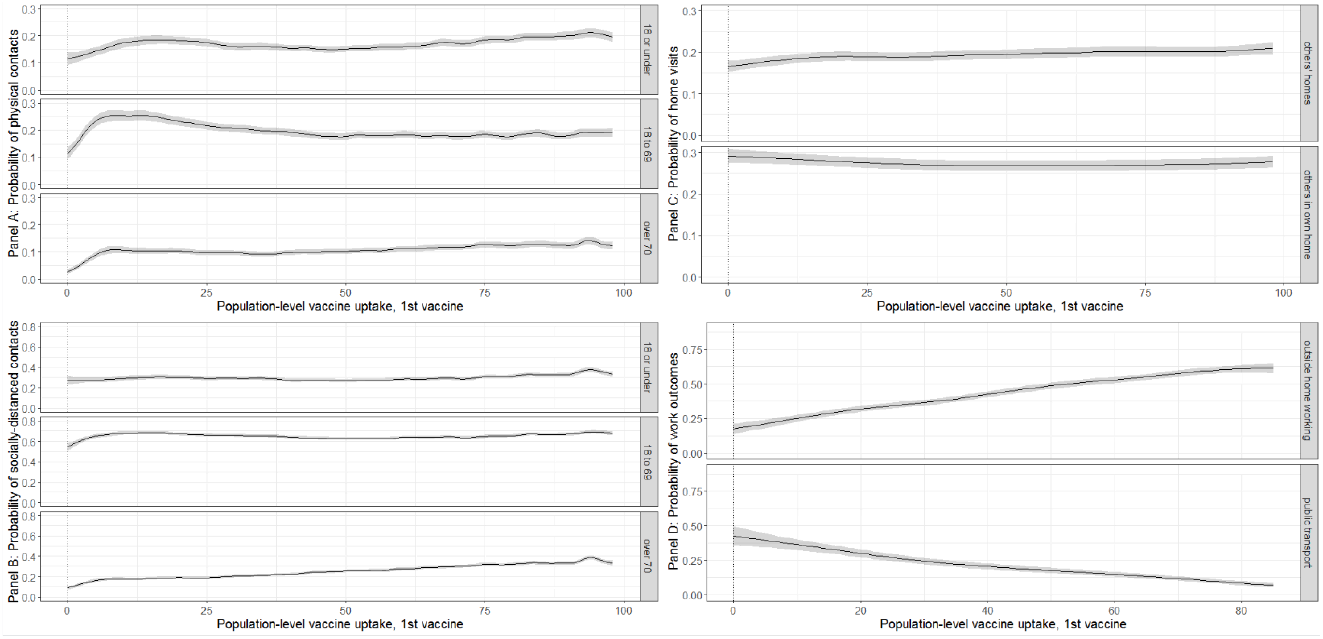
Probabilities of behavioural outcomes by population level vaccination %, first dose. Top left (panel A): past 7-day reported physical, outside of household contacts; bottom left (Panel B): past 7-day reported socially-distanced, outside of household contacts; top right (Panel C): past 7-day reported home visits; bottom right (Panel D): past 7-day reported work outcomes for those that are working or in education. “18 or under”, “18 to 69” and “over 70” denote the ages of the people with whom that individuals in the sample had contact.

The probability of any physical contacts outside of the household (with under-18s, 18 to 69 year-olds, and over-70s) increased as population-level vaccination coverage increased overall (Fig 4A). For all three contact groups, there were distinct increases in the probability of any physical contacts when the population vaccination levels were between 5% and 25%, when the most vulnerable individuals were being vaccinated. This is illustrated in Fig 5, where the rate of vaccination of over-70s is plotted against the rate of vaccination among the under-70s for the UK as a whole. On average, by the time 10% of under-70s had received a first vaccination, approximately 80% of over-70s had received their first vaccination. For physical contacts with those 18 to 69 years, the probability of reporting any physical contacts outside of the household in the past 7 days reached a peak of around 0.3, which afterwards stabilized at a probability of approximately 0.2. The probability of any physical contact with over 70s was initially almost zero until 5% of the population received their first vaccination, after which the probability of physical contacts with this age-group started to increase rapidly to 0.1-0.15. Increases were also observed for physical contacts with under-18s when population-level vaccination uptake increased from 0-12.5%.

**Figure 5:**
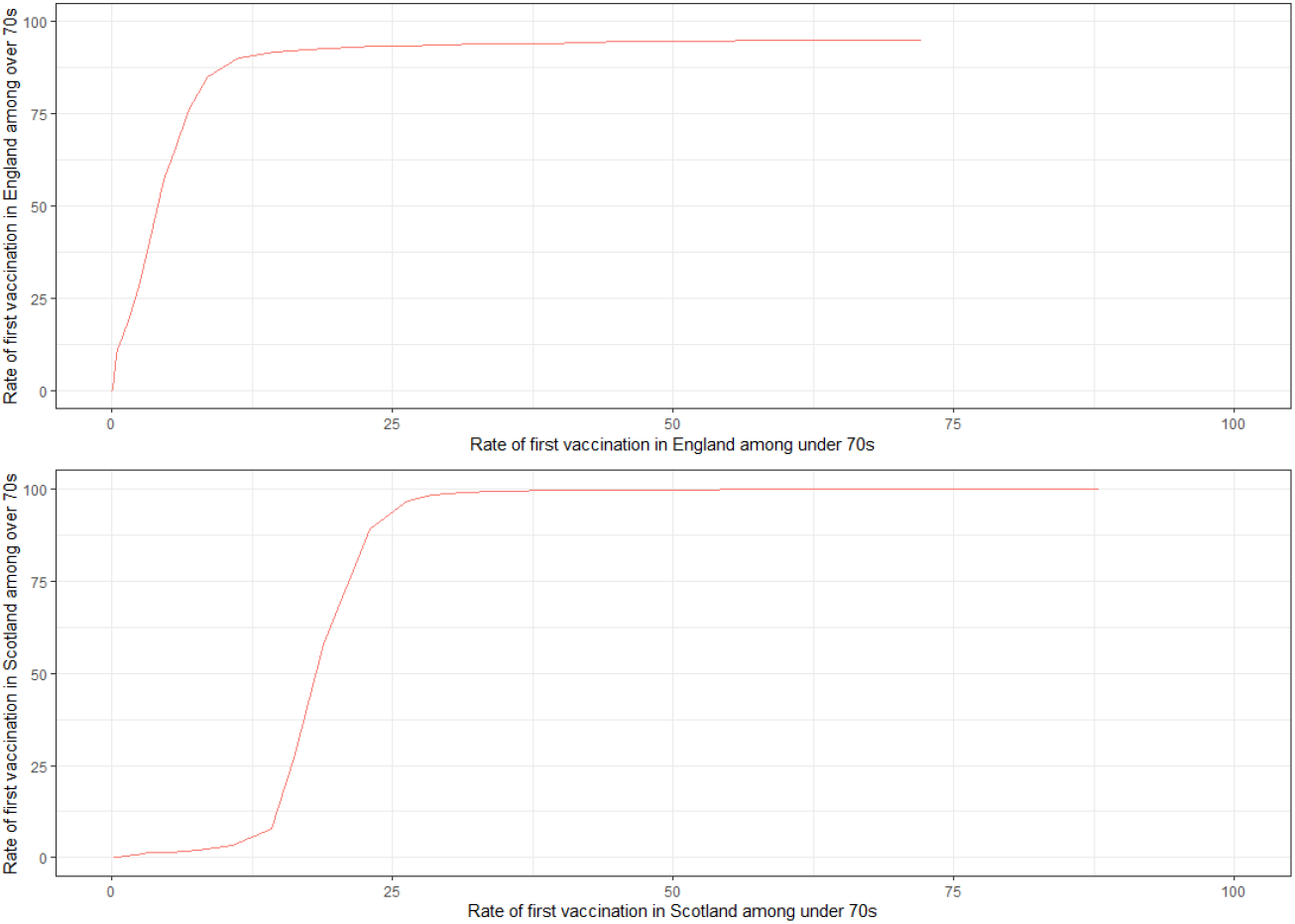
Vaccination rate among over-70s in England and Scotland versus vaccination rates in younger adults (source: https://coronavirus.data.gov.uk/details/download). Data were unavailable for Northern Ireland and Wales.

The trends for socially-distanced contacts outside of the household with under-18s and 18 to 69 year-olds were reasonably flat (Figure 4B), with the probability of any contacts remaining at around 0.3 and 0.7, respectively. For socially-distanced contacts outside of the household with over 70s, the probability of having any contacts increased steadily with population-level vaccination, peaking at around 0.4 when most eligible individuals were vaccinated.

Increases in population-level vaccination uptake from 0 to 25% were associated with a small gradual increase in the probability of visiting others’ homes (from 0.17 [95%CI: 0.16 to 0.18] to 0.21 [95%CI: 0.19 to 0.22], Fig 4C). No meaningful difference in the association between the probability of others’ visits to own home and population-level vaccination was observed.

The probability of working outside of the home (note this is restricted to those who reported that they are currently working) increased concurrently with increasing population-level vaccination uptake from around 0.18 [95%CI: 0.14 to 0.21] to 0.62 [95%CI: 0.58 to 0.65] (Fig 4D). The probability of using public transport for work travel (also restricted to those who reported that they are currently working) in the past 7 days declined concurrently with increasing population-level vaccination uptake 0.43 [95%CI: 0.36 to 0.50] to 0.07 [95%CI: 0.05 to 0.09] (Fig 4D). A lower probability of taking public transport could be explained by those that increasingly return to outside the home working using private, rather than public transport.

## Discussion

We evaluated how behaviour, measured by the probabilities of physical or socially-distanced contacts with individuals outside the home, visits to others’ homes or of others to one’s own home, work location, and use of public transport, varied in the UK: (i) over time since first vaccination among adults aged 18 to 64; (ii) over time since the last vulnerable person in the household having their first vaccination among unvaccinated adults aged 18 to 64 without long-term health conditions; and (iii) by local population-level vaccine uptake among individuals aged 2 to 95 years old. There appeared to be a general increase in the probability of socialising, in terms of physical contacts, socially-distanced contacts, and working outside the home over time since first vaccination, especially once high-risk individuals were vaccinated. However, we found little to no response in the trends of the measured behaviours to the vaccination itself. There was also limited evidence to suggest that unvaccinated individuals adjusted their behaviour in response to all vulnerable people in their households being vaccinated. There were, however, increases in the population’s probabilities of contacts, in terms of physical contacts, socially-distanced contacts, and working outside the home as population vaccination rates increased. More specifically, a surge in physical contacts, especially for contacts with those aged 70 years and older, and to a lesser extent socially-distanced contacts, was reported when people that were most vulnerable had their first vaccination. Contacts, home visits, and working outside the home increased steadily as vaccination rates in the population increased. This was the case even after controlling for region-specific time trends (appendix 7), which are expected to capture factors such as public health messaging and restrictions which are also related to the outcomes. This suggests risk compensatory behaviours related to population level vaccination beyond policy changes.

Strengths of this study include the large, nationally-representative, random sample of individuals from the UK. The wide range of behavioural outcomes allows for insights into behavioural change, and internal validity in observing consistent patterns of behaviour where expected. That we observed behaviours under various changing public health mitigation measures is useful for making more general statements about behaviour than would be permissible if we had only observed individuals in strict lockdown or under constant conditions. The data that were collected allow us to control for a set of potentially confounding variables, including household composition, individual characteristics, sociodemographic variables, and time.

Limitations include that the survey is based on self-reported behaviours. Individuals tend to underreport socially undesirable/stigmatizing behaviours^22^, e.g. having many contacts during periods of strict lockdown, which could translate to reported vaccination behaviours. However, respondents were advised that their answers would be confidential and that the results from the survey would influence the government’s response to the pandemic, the survey was administered by the national statistics agency in the UK, and individuals were paid to participate. These factors lessen underreporting incentives^23-25^. Underreporting would lead to downward bias on estimates; for example, if individuals reported fewer physical contacts than they actually had, then our estimated probability of having these contacts would be lower than the true probability. The fact that only 36% reported having a physical contact in the week they received the vaccination – an increase of 10% compared to the week before the vaccination - suggests that some respondents either thought a vaccination appointment should not be counted as a physical contact or that they perhaps generally underreported contacts. Given this, we consider these results to be lower bounds of risk-compensatory behaviour. There are other behaviours that were not captured in these outcomes. For example, we did not capture non-work travel (e.g. trips made inter-/intra-regionally). Moreover, we did not account for the risk of the types of interaction in these outcomes (e.g. high risk contact in nightclubs vs low risk attending GP to have a vaccine dose, or high risk contact working in a public-facing role compared to working in a socially-distanced office). Finally, due to limited memory in the secure computing environment, it was not possible to estimate models with individual-specific random effects.

Previous literature about risk compensation associated with SARS-CoV-2 vaccination has been conflicting, with some reporting in favour of behavioural change^18-19^ and others against^17^, but in either case, these were merely conjecture. Our evidence is in favour of a limited degree of risk compensation, though this is related more to population vaccination level, rather than individual level vaccination. Physical contacts, socially-distanced contacts, and working outside the home were positively related to population level vaccination. These behaviours are in turn positively related to transmission. This is of concern if individuals are less cautious given evidence of waning immunity^11-12^, especially if vulnerable individuals have not yet received booster vaccinations. Thus, understanding this behaviour may be of further importance as booster vaccinations are administered^26^.

We provide the first evidence on the behavioural response to COVID-19 vaccination. Patterns of behaviour did not respond to individual or household vaccinations, but did respond to population-level vaccination. This evidence suggests that population-rate-based risk compensation may have pervaded behaviours in the UK during the SARS-CoV-2 pandemic.

## Methods

### Data

Data are taken from the UK Office for National Statistics Coronavirus (COVID-19) Infection Survey (ISRCTN21086382; https://www.ndm.ox.ac.uk/covid-19/covid-19-infection-survey/protocol-and-information-sheets; details in Pouwels et al., 2021^27^). Individuals were approached from households that were randomly selected from previous surveys and address lists in England, Northern Ireland, Scotland, and Wales to provide a representative sample of the UK population. The survey consists of repeated cross-sectional household surveys with additional serial sampling and longitudinal follow-up. Data collected between 1^st^ October 2020 and 15^th^ September 2021 were used for this analysis. Ethical approval was granted by South Central–Berkshire B Research Ethics Committee (20/SC/0195). Appendix 1 provides further details of the sampling design.

Data include information on the behavioural outcomes of interest, sociodemographic characteristics, and medical information (https://www.ndm.ox.ac.uk/covid-19/covid-19-infection-survey/case-record-forms for survey questionnaires).

### Vaccination status and population-level uptake

Patients were asked about their vaccination status, including type, number of doses and date(s). Survey participants from England were also linked to administrative records from the National Immunisation Management Service (NIMS)^27^. Where available, we used records from NIMS, otherwise we used records from the survey, since linkage was periodic and NIMS does not contain information about vaccinations received abroad or in Northern Ireland, Scotland, or Wales. Where records were available in both, agreement on type was 98% and dates 95% within ±7 days.

Data on population level uptake were taken from publicly available government sources (https://coronavirus.data.gov.uk/details/download). This was mapped to the participants’ data at the level of CIS geographical regions, which is 133 geographical areas within the four nations on which CIS collects data.

### Outcomes

10 behavioural outcomes were self-reported by individuals:

i. The number of physical contacts, e.g. handshake, personal care, including with personal protective equipment, with individuals aged <18 years old in the past 7 days;
ii. The number of physical contacts with individuals aged 18-69 years old in the past 7 days;
iii. The number of physical contacts with individuals aged 70 years and over in the past 7 days;
iv. The number of socially distanced contacts, worded as, “direct, but not physical, contact”, with individuals aged <18 years old in the past 7 days;
v. The number of socially distanced contacts with individuals aged 18-69 years old in the past 7 days;
vi. The number of socially distanced contacts with individuals aged 70 years and over in the past 7 days;
vii. The number of times the participant spend one hour or longer inside their own home with someone from another household in the past 7 days;
viii. The number of times the participant spend time one hour or longer inside the building of another person’s home in the past 7 days;
ix. Among those that reported engagement in work or studying: mode of travel to work/place of education (grouped as public transport versus other for the current analyses) in the past 7 days; and
x. Among those that reported engagement in work or studying: main work/study location in the past 7 days.

The physical and socially distanced contact variables (i – vi) were recorded as ordered variables with 5 response categories (0, 1-5, 6-10, 11-20, 21 or more). Both types of home visits (vii and viii) were recorded as ordered variables with 8 response categories (0, 1, 2, 3, 4, 5, 6, 7 times or more). Work or study location was recorded as an ordered variable with three categories (working from home, working from a mix of home and outside of home, working from outside the home).

### Sample eligibility

#### Behaviour changes among vaccinated individuals

Individuals that met the following criteria were included: age 18 years or older, or an individual self-reported a long term health condition (in the UK those with underlying health conditions aged 16 years and over were prioritised after those aged 65 years and over). Preliminary testing plotting difference of the smooths^28^ indicated two groups, patient-facing healthcare workers and over 65s had different behaviours to those under 65 or that did not work in healthcare settings; results are presented in the appendix 2. Patient-facing healthcare workers were both prioritised for vaccination and also are expected to have had unusually high contacts during periods of strict mitigation measures given the nature of their job. Older individuals have much higher risk of severe outcomes, generally less frequent contact patterns, and an increasingly small number of unvaccinated older individuals remaining unvaccinated^12^. Individuals were only included if there was evidence that they were vaccinated at any point during the survey to avoid that those not willing to get vaccinated, who are expected to have different behaviours than those that are willing to get vaccinated.

Individuals were included if they had at least one pre-vaccination and one post-vaccination observation between 1^st^ October 2020 and 15^th^ September 2021.

#### Influence of vulnerable household member vaccination

This analysis included individuals that lived in a household with a vulnerable individual (defined as either >65 years or having a long term health condition) but themselves were not considered vulnerable and not yet vaccinated, with visits from the 1^st^ October 2020 and 15^th^ September 2021.

#### Influence of population-level uptake

All individuals with visits in the period 1^st^ October 2020 and 15^th^ September 2021 were included in analyses, regardless of vaccination status or reported job.

### Statistical analyses

All analyses used generalized additive ordered categorical regressions that were estimated on the ordinal outcomes^29^. In these models, the linear predictor is a latent variable with estimated thresholds that mark the transitions between levels of the ordered categorical response. For public transport for working travel, which as a binary outcome, a generalized additive logistic regression was used.

Predicted probabilities were estimated for all response categories of the ordinal outcomes. We then collapsed these into binary variables, where the values of the outcomes at zero are treated as zero, and one otherwise (posterior probabilities of being in each category, and their summation to construct the reported probability, are provided in the appendix 3 for physical, outside of household contacts with 18 to 69 year-olds). In this way, the binary variable captures the margin between “none” and “any”: for contacts, this variable is either no outside of household contacts in the past 7 days (0) or any outside of household contacts in the past 7 days (1); for home visits it is either no visits in the past 7 days (0) or any home visits in the past 7 days (1); for work location, this is either working at home every day in the past 7 days (0) or working in outside of home location (i.e. office, café, etc.) one or more times in the past 7 days (1); and for mode of travel to work, this is either private transport in the past 7 days (0) or public transport (1). Using the ordinal variables in models allows us to exploit all of the information in the data, and collapsing them into posterior binary variables eases exposition, aligns the results across outcomes with different numbers of categories, and focusses the analyses on the margin of “none” and “any” which is most useful for understanding transmission.

#### Behaviour changes among vaccinated individuals

This analysis examined the behavioural response to individual vaccination. Outcomes were modelled from 180 days prior to first vaccination to 120 after the first vaccination (given few visits outside of this range). To capture non-linear behavioural change, we used thin plate splines on time from first vaccination (the number of basis functions (*k* in the mgcv package^30^) was the number of unique values divided by 3). The degree of smoothing of the splines was optimised using a fast implementation of restricted maximum likelihood (REML)^30,31^. Regressions controlled for confounding with individual-specific characteristics of gender, ethnicity, socioeconomic status (rank of index of multiple deprivation, calculated separately for each country in the United Kingdom^32-35^, geographical region, urban/rural classification^36-38^, the stage of lockdown (and corresponding restrictions applied), household size, whether the household is multigenerational (household individuals aged 16 or younger and individuals aged school year 12 to age 49 and individuals aged 50+), and if the individual ever reported a long-term healthcare condition. Calendar time by region/country (9 regions in England and Northern Ireland, Scotland, and Wales) interactions were implemented to control for non-linear confounding of region/country-specific behavioural change over time (see appendix 7 for an example of physical contacts with under 18 year-olds). Thin plate splines were also applied to age to control for non-linear confounding of age-specific behaviour.

Testing changes in behavioural responses to individual vaccination involved testing trends pre- and post-vaccination. More specifically, the gradient in the outcomes (over time from vaccination) in a period before vaccination (−50 days to −14 days) was tested against two periods (1 to 13 days and 14 to 50 days) after vaccination, arguing that longer-term changes were less likely related to vaccination. This was conducted using 10,000 simulations based on pseudo-random draws from the Bayesian posterior distributions of the fitted GAM models using the Gaussian approximation to the posterior of the model coefficients. The pseudo-random draws are obtained from a multivariate normal with mean vector equal to the estimated model coefficients and covariance matrix equal to the covariance matrix of the coefficients. For each draw, the gradient of the outcome was computed for the pre- and the two post-periods; and the differences in the gradients between the pre-period and each of the post-periods, respectively, were taken. Across the draws, the median, 2.5^th^, and 97.5^th^ percentiles of the distribution of differences were taken and compared to assess if the gradients of the outcomes changed between pre- and post-vaccination periods.

#### Influence of vulnerable household member vaccination

This analysis examined the behavioural response to vaccination of vulnerable household members as described above. This analysis was similar to that for vaccinated individuals, except the definition of vaccination date was altered to be the date of vaccination for the last vulnerable individual in the household to be vaccinated. In addition, this analysis was limited to the subset of (a) individuals considered to be non-vulnerable and (c) observations until individuals themselves received a first vaccination. To capture non-linear behavioural change, we used thin plate splines on time from first household vaccination^39^ (the number of basis functions (*k* in the mgcv package^30^) was based on the number of unique values divided by 3). Model specification was otherwise identical to the analysis of vaccinated individuals.

#### Influence of population-level uptake

This analysis examined the behavioural response to vaccination rates at the population level rather than to individual vaccination. In these models, the time from first vaccination variable was replaced with the percentage of individuals in the population that had been vaccinated. Daily rates of vaccine uptake were available for local areas and these were merged to the CIS data by CIS geographical area to enable modelling. To capture non-linear behavioural change, we used thin plate splines on population-level-vaccination^39^ (the number of basis functions (*k* in the mgcv package^30^) was the number of unique values divided by 3). In addition, a variable for whether individuals were patient-facing healthcare workers was included. Thin plate splines were also applied to age to control for non-linear confounding of age-specific behaviour (where again the number of basis functions was the number of unique values divided by 3); age was truncated at 95 to reduce the influence of outliers. A categorical variable was defined which indicated if the individual was a child, a vaccinated adult or an unvaccinated adult. This variable was interacted with the thin-plate splines on population level vaccination to allow for differential behavioural responses for each of the groups.

#### Specification and sensitivity analyses

The proportional odds assumption was tested in preliminary analyses by interacting the threshold parameters with a post-vaccination dummy variable. No evidence of variation in the response over the range of the outcomes was found. We tested tensor splines for time-age-region interactions (knots divided the time range at 3-day intervals and the age range at 3-year intervals) to control for non-linear confounding of age- and region-specific behavioural change over time. There was no improvement in model fit nor impact on the difference in the outcomes over time. Thus, the simpler specification, i.e. region by time interaction, was retained.

Models examined if behaviours were consistent in three groups (relative to the remaining sample): those over age 65, reporting working in patient-facing healthcare, and reporting a long-term health condition. Evidence of different patterns of behaviour were found for those over 65 years and for patient-facing healthcare workers, but not for those self-reporting long-term health conditions (results are presented in the appendix 2). Differential behavioural trends in response to ChAdOx1 versus BNT162b2 vaccines were evaluated by plotting difference of the smooths (other types of vaccine, e.g. mRNA-1273 (Moderna), were controlled for separately). No differences were observed in behaviours for ChAdOx1 versus the BNT162b2 vaccines (appendix 4). For the population-level vaccination models, including an interaction between the thin-plate splines for time-from-vaccination and a categorical variable indicating if the individual was a child, a vaccinated adult or an unvaccinated adult improved the fit of the model and was therefore used. A model that included both time from own vaccination and population level vaccination were estimated to ensure that the results were robust.

Models that use the date of second, rather than first, vaccination yielded similar patterns across the outcomes. Results are shown in appendix 5. In addition, we analysed responses to population level vaccination among those that had been vaccinated and consistent with the eligibility criteria described for the analysis of own vaccination. Results are presented in appendix 6. In contrast to the full sample, an initial peak in the probability of contacts in the first 25% of population vaccination is not observed, suggesting this peak is driven by children and the unvaccinated; otherwise the patterns of behaviour are consistent.

## Data Availability

De-identified study data are available for access by accredited researchers in the ONS Secure Research Service (SRS) for accredited research purposes under part 5, chapter 5 of the Digital Economy Act 2017.

## Data Availability

Data are still being collected for the COVID-19 Infection Survey. De-identified study data are available for access by accredited researchers in the ONS Secure Research Service (SRS) for accredited research purposes under part 5, chapter 5 of the Digital Economy Act 2017. For further information about accreditation, contact or visit the SRS website.

## Acknowledgements

This COVID-19 Infection Survey is funded by the Department of Health and Social Care with in-kind support from the Welsh Government, the Department of Health on behalf of the Northern Ireland Government and the Scottish Government. JB is also supported by the NIHR Oxford Biomedical Research Centre. KBP and ASW are supported by the National Institute for Health Research Health Protection Research Unit (NIHR HPRU) in Healthcare Associated Infections and Antimicrobial Resistance at the University of Oxford in partnership with Public Health England (PHE) (NIHR200915). ASW is also supported by the NIHR Oxford Biomedical Research Centre. KBP is also supported by the Huo Family Foundation. ASW is also supported by core support from the Medical Research Council UK to the MRC Clinical Trials Unit [MC_UU_12023/22] and is an NIHR Senior Investigator. PCM is funded by Wellcome (intermediate fellowship, grant ref 110110/Z/15/Z) and holds an NIHR Oxford BRC Senior Fellowship award. The views expressed are those of the authors and not necessarily those of the National Health Service, NIHR, Department of Health, or PHE. The funder/sponsor did not have any role in the design and conduct of the study; collection, management, analysis, and interpretation of the data; preparation, review, or approval of the manuscript; and decision to submit the manuscript for publication. All authors had full access to all data analysis outputs (reports and tables) and take responsibility for their integrity and accuracy.

We are grateful for the support of all COVID-19 Infection Survey participants and the COVID-19 Infection Survey team:

Office for National Statistics: Sir Ian Diamond, Emma Rourke, Ruth Studley, Tina Thomas, Duncan Cook.

Office for National Statistics COVID Infection Survey Analysis and Operations teams, in particular Daniel Ayoubkhani, Russell Black, Antonio Felton, Megan Crees, Joel Jones, Lina Lloyd, Esther Sutherland.

University of Oxford, Nuffield Department of Medicine: Ann Sarah Walker, Derrick Crook, Philippa C Matthews, Tim Peto, Emma Pritchard, Nicole Stoesser, Karina-Doris Vihta, Jia Wei, Alison Howarth, George Doherty, James Kavanagh, Kevin K Chau, Stephanie B Hatch, Daniel Ebner, Lucas Martins Ferreira, Thomas Christott, Brian D Marsden, Wanwisa Dejnirattisai, Juthathip Mongkolsapaya, Sarah Cameron, Phoebe Tamblin-Hopper, Magda Wolna, Rachael Brown, Sarah Hoosdally, Richard Cornall, Yvonne Jones, David I Stuart, Gavin Screaton.

University of Oxford, Nuffield Department of Population Health: Koen Pouwels.

University of Oxford, Big Data Institute: David W Eyre, Katrina Lythgoe, David Bonsall, Tanya Golubchik, Helen Fryer.

University of Oxford, Radcliffe Department of Medicine: John Bell.

Oxford University Hospitals NHS Foundation Trust: Stuart Cox, Kevin Paddon, Tim James. University of Manchester: Thomas House.

Public Health England: John Newton, Julie Robotham, Paul Birrell.

IQVIA: Helena Jordan, Tim Sheppard, Graham Athey, Dan Moody, Leigh Curry, Pamela Brereton.

National Biocentre: Ian Jarvis, Anna Godsmark, George Morris, Bobby Mallick, Phil Eeles.

Glasgow Lighthouse Laboratory: Jodie Hay, Harper VanSteenhouse.

Department of Health and Social Care: Jessica Lee.

Welsh Government: Sean White, Tim Evans, Lisa Bloemberg.

Scottish Government: Katie Allison, Anouska Pandya, Sophie Davis.

Public Health Scotland: David I Conway, Margaret MacLeod, Chris Cunningham.

