## Appendices for "COVID-19 vaccination, risk-compensatory behaviours, and contacts in the UK"

Appendix 1: Sampling design

The following information on the sampling design can also be found at: <https://www.ons.gov.uk/>peoplepopulationandcommunity/healthandsocialcare/conditionsanddiseases/methodologies/covid19infectionsurveypilotmethodsandfurtherinformation#study-design-sampling

At the start of the study at the end of April 2020, the sample for the survey was drawn mainly from the Annual Population Survey (APS), which consists collectively of those who successfully completed the last wave of the Labour Force Survey (LFS) or local LFS boost, and who had consented to future contact regarding research.

Around 38,000 households respond to the LFS each quarter and it is the largest regular household survey in the UK. The sampling frame for the LFS is the Postal Address File of small users, which contains approximately 26 million addresses. Only private households are included in the sample. People living in care homes, other communal establishments and hospitals are not included.

At the start of the study, all respondents to the COVID-19 Infection Survey were individuals who have previously participated in an Office for National Statistics (ONS) social survey, which means the number of ineligible addresses in the sample is substantially reduced. To take part, invited households opted into the survey by contacting IQVIA, a company working on behalf of the ONS, to arrange a visit.

Since the end of July 2020, we further expanded the survey to invite a random sample of households from AddressBase, which is a commercially available list of addresses maintained by the Ordnance Survey.

In August 2020, we further expanded the study with the aim of increasing from 28,000 people tested per fortnight in England to 150,000 people tested per fortnight by October 2020 until March 2022. A random sample of households from AddressBase was invited for this expansion.

Appendix 2: age, long term health conditions, and health and social care workers testing

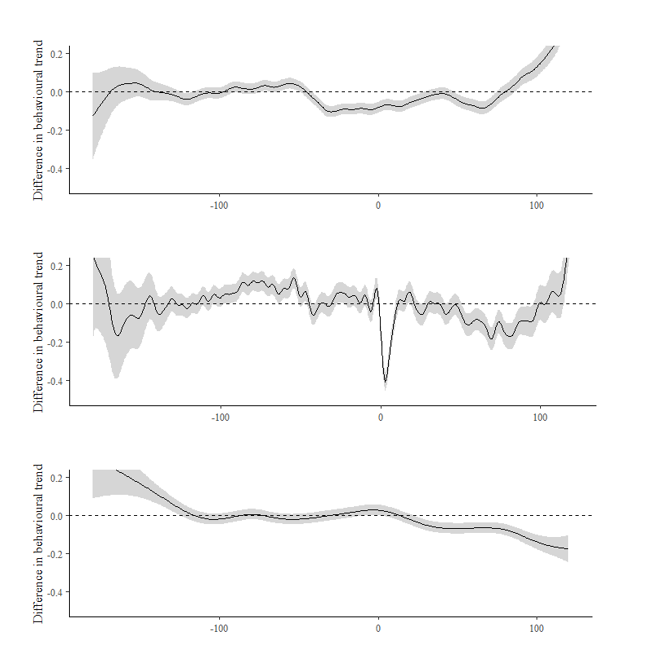

Table A1: differences in smooths for age>65 (vs. age<=65). Top panel: others in own home in the past 7 days. Middle panel: physical contacts with 18 to 69 year-olds in the past 7 days. Bottom panel: Socially-distanced contacts with 18 to 69 year-olds in the past 7 days. Evidence to suggest behavioural trends diverge between ages over and under 65, hence main analyses focussed on those aged 18-64.

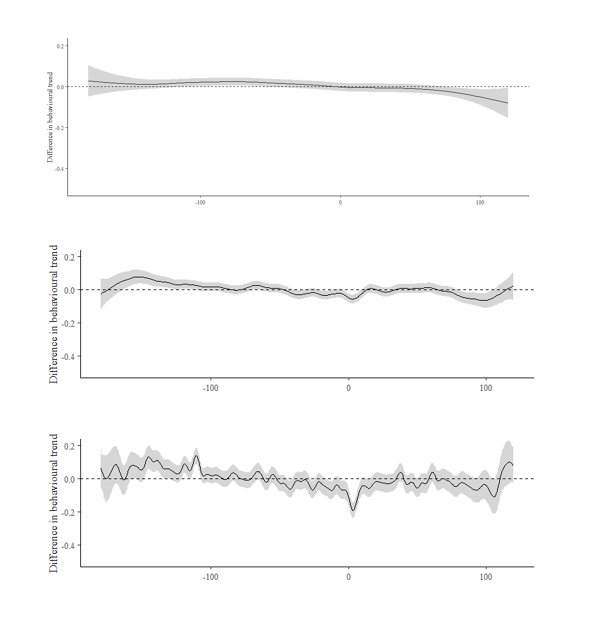

Table A2: differences in smooths for individuals with long-term health conditions (vs. individuals without long-term health conditions). Top panel: others in own home in the past 7 days. Middle panel: physical contacts with 18 to 69 year-olds in the past 7 days. Bottom panel: Socially-distanced contacts with 18 to 69 year-olds in the past 7 days. No evidence to suggest behavioural trends diverge between individuals with and without long-term health conditions.

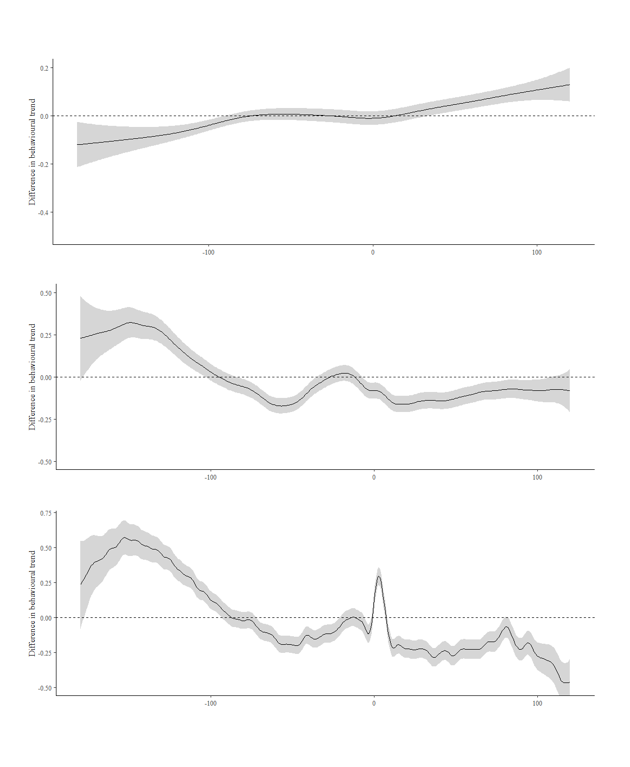

Table A3: differences in smooths for patient-facing healthcare workers (vs. non-health and social care workers). Top panel: others in own home in the past 7 days. Middle panel: physical contacts with 18 to 69 year-olds in the past 7 days. Bottom panel: Socially-distanced contacts with 18 to 69 year-olds in the past 7 days. Evidence to suggest behavioural trends diverge between individuals who are and are not health and social care workers.

Appendix 3: prediction for levels of ordered variable and construction of probability of any 18 to 69 year old, outside of household, physical contacts

| Time from vaccination | fit.1 | fit.2 | fit.3 | fit.4 | fit.5 | Probability of some contacts |
| --- | --- | --- | --- | --- | --- | --- |
| -180 | 0.871 | 0.111 | 0.010 | 0.004 | 0.004 | 0.129 |
| -179 | 0.869 | 0.113 | 0.010 | 0.004 | 0.004 | 0.131 |
| -178 | 0.867 | 0.115 | 0.010 | 0.004 | 0.004 | 0.133 |
| -177 | 0.865 | 0.116 | 0.010 | 0.004 | 0.004 | 0.135 |
| -176 | 0.864 | 0.118 | 0.011 | 0.004 | 0.004 | 0.136 |
| -175 | 0.863 | 0.119 | 0.011 | 0.004 | 0.004 | 0.137 |
| -174 | 0.862 | 0.119 | 0.011 | 0.004 | 0.004 | 0.138 |
| -173 | 0.861 | 0.120 | 0.011 | 0.004 | 0.004 | 0.139 |
| -172 | 0.861 | 0.120 | 0.011 | 0.004 | 0.004 | 0.139 |
| -171 | 0.861 | 0.120 | 0.011 | 0.004 | 0.004 | 0.139 |
| -170 | 0.860 | 0.121 | 0.011 | 0.004 | 0.004 | 0.140 |
| -169 | 0.860 | 0.121 | 0.011 | 0.004 | 0.004 | 0.140 |
| -168 | 0.860 | 0.121 | 0.011 | 0.004 | 0.004 | 0.140 |
| -167 | 0.859 | 0.122 | 0.011 | 0.004 | 0.004 | 0.141 |
| -166 | 0.859 | 0.122 | 0.011 | 0.004 | 0.004 | 0.141 |
| -165 | 0.858 | 0.122 | 0.011 | 0.004 | 0.004 | 0.142 |
| -164 | 0.858 | 0.122 | 0.011 | 0.004 | 0.004 | 0.142 |
| -163 | 0.858 | 0.122 | 0.011 | 0.004 | 0.004 | 0.142 |
| -162 | 0.858 | 0.122 | 0.011 | 0.004 | 0.004 | 0.142 |
| -161 | 0.859 | 0.122 | 0.011 | 0.004 | 0.004 | 0.141 |
| -160 | 0.859 | 0.122 | 0.011 | 0.004 | 0.004 | 0.141 |
| -159 | 0.859 | 0.122 | 0.011 | 0.004 | 0.004 | 0.141 |
| -158 | 0.858 | 0.122 | 0.011 | 0.004 | 0.004 | 0.142 |
| -157 | 0.858 | 0.123 | 0.011 | 0.004 | 0.004 | 0.142 |
| -156 | 0.858 | 0.123 | 0.011 | 0.004 | 0.004 | 0.142 |
| -155 | 0.858 | 0.123 | 0.011 | 0.004 | 0.004 | 0.142 |
| -154 | 0.857 | 0.123 | 0.011 | 0.004 | 0.004 | 0.143 |
| -153 | 0.858 | 0.123 | 0.011 | 0.004 | 0.004 | 0.142 |
| -152 | 0.858 | 0.123 | 0.011 | 0.004 | 0.004 | 0.142 |
| -151 | 0.859 | 0.122 | 0.011 | 0.004 | 0.004 | 0.141 |
| -150 | 0.859 | 0.122 | 0.011 | 0.004 | 0.004 | 0.141 |
| -149 | 0.859 | 0.122 | 0.011 | 0.004 | 0.004 | 0.141 |
| -148 | 0.859 | 0.122 | 0.011 | 0.004 | 0.004 | 0.141 |
| -147 | 0.859 | 0.122 | 0.011 | 0.004 | 0.004 | 0.141 |
| -146 | 0.858 | 0.122 | 0.011 | 0.004 | 0.004 | 0.142 |
| -145 | 0.858 | 0.122 | 0.011 | 0.004 | 0.004 | 0.142 |
| -144 | 0.859 | 0.122 | 0.011 | 0.004 | 0.004 | 0.141 |
| -143 | 0.859 | 0.121 | 0.011 | 0.004 | 0.004 | 0.141 |
| -142 | 0.860 | 0.121 | 0.011 | 0.004 | 0.004 | 0.140 |
| -141 | 0.860 | 0.121 | 0.011 | 0.004 | 0.004 | 0.140 |
| -140 | 0.859 | 0.122 | 0.011 | 0.004 | 0.004 | 0.141 |
| -139 | 0.859 | 0.122 | 0.011 | 0.004 | 0.004 | 0.141 |
| -138 | 0.858 | 0.122 | 0.011 | 0.004 | 0.004 | 0.142 |
| -137 | 0.858 | 0.122 | 0.011 | 0.004 | 0.004 | 0.142 |
| -136 | 0.858 | 0.122 | 0.011 | 0.004 | 0.004 | 0.142 |
| -135 | 0.859 | 0.122 | 0.011 | 0.004 | 0.004 | 0.141 |
| -134 | 0.859 | 0.122 | 0.011 | 0.004 | 0.004 | 0.141 |
| -133 | 0.859 | 0.122 | 0.011 | 0.004 | 0.004 | 0.141 |
| -132 | 0.859 | 0.122 | 0.011 | 0.004 | 0.004 | 0.141 |
| -131 | 0.859 | 0.122 | 0.011 | 0.004 | 0.004 | 0.141 |
| -130 | 0.858 | 0.122 | 0.011 | 0.004 | 0.004 | 0.142 |
| -129 | 0.858 | 0.123 | 0.011 | 0.004 | 0.004 | 0.142 |
| -128 | 0.857 | 0.123 | 0.011 | 0.004 | 0.004 | 0.143 |
| -127 | 0.857 | 0.124 | 0.011 | 0.004 | 0.004 | 0.143 |
| -126 | 0.856 | 0.124 | 0.011 | 0.004 | 0.004 | 0.144 |
| -125 | 0.856 | 0.124 | 0.011 | 0.004 | 0.004 | 0.144 |
| -124 | 0.857 | 0.124 | 0.011 | 0.004 | 0.004 | 0.143 |
| -123 | 0.857 | 0.123 | 0.011 | 0.004 | 0.004 | 0.143 |
| -122 | 0.858 | 0.123 | 0.011 | 0.004 | 0.004 | 0.142 |
| -121 | 0.858 | 0.123 | 0.011 | 0.004 | 0.004 | 0.142 |
| -120 | 0.857 | 0.123 | 0.011 | 0.004 | 0.004 | 0.143 |
| -119 | 0.857 | 0.124 | 0.011 | 0.004 | 0.004 | 0.143 |
| -118 | 0.856 | 0.124 | 0.011 | 0.004 | 0.004 | 0.144 |
| -117 | 0.855 | 0.125 | 0.011 | 0.004 | 0.004 | 0.145 |
| -116 | 0.854 | 0.126 | 0.011 | 0.004 | 0.004 | 0.146 |
| -115 | 0.853 | 0.126 | 0.011 | 0.004 | 0.004 | 0.147 |
| -114 | 0.852 | 0.127 | 0.012 | 0.004 | 0.004 | 0.148 |
| -113 | 0.851 | 0.128 | 0.012 | 0.004 | 0.005 | 0.149 |
| -112 | 0.850 | 0.129 | 0.012 | 0.004 | 0.005 | 0.150 |
| -111 | 0.849 | 0.130 | 0.012 | 0.004 | 0.005 | 0.151 |
| -110 | 0.849 | 0.130 | 0.012 | 0.004 | 0.005 | 0.151 |
| -109 | 0.849 | 0.131 | 0.012 | 0.004 | 0.005 | 0.151 |
| -108 | 0.848 | 0.131 | 0.012 | 0.004 | 0.005 | 0.152 |
| -107 | 0.848 | 0.131 | 0.012 | 0.004 | 0.005 | 0.152 |
| -106 | 0.848 | 0.131 | 0.012 | 0.004 | 0.005 | 0.152 |
| -105 | 0.847 | 0.132 | 0.012 | 0.004 | 0.005 | 0.153 |
| -104 | 0.847 | 0.132 | 0.012 | 0.004 | 0.005 | 0.153 |
| -103 | 0.847 | 0.132 | 0.012 | 0.004 | 0.005 | 0.153 |
| -102 | 0.847 | 0.132 | 0.012 | 0.004 | 0.005 | 0.153 |
| -101 | 0.847 | 0.132 | 0.012 | 0.004 | 0.005 | 0.153 |
| -100 | 0.847 | 0.132 | 0.012 | 0.004 | 0.005 | 0.153 |
| -99 | 0.847 | 0.132 | 0.012 | 0.004 | 0.005 | 0.153 |
| -98 | 0.846 | 0.132 | 0.012 | 0.004 | 0.005 | 0.154 |
| -97 | 0.846 | 0.133 | 0.012 | 0.004 | 0.005 | 0.154 |
| -96 | 0.845 | 0.133 | 0.012 | 0.004 | 0.005 | 0.155 |
| -95 | 0.845 | 0.134 | 0.012 | 0.004 | 0.005 | 0.155 |
| -94 | 0.844 | 0.134 | 0.012 | 0.004 | 0.005 | 0.156 |
| -93 | 0.844 | 0.135 | 0.012 | 0.004 | 0.005 | 0.156 |
| -92 | 0.843 | 0.135 | 0.012 | 0.004 | 0.005 | 0.157 |
| -91 | 0.843 | 0.135 | 0.012 | 0.004 | 0.005 | 0.157 |
| -90 | 0.843 | 0.135 | 0.012 | 0.004 | 0.005 | 0.157 |
| -89 | 0.843 | 0.135 | 0.012 | 0.004 | 0.005 | 0.157 |
| -88 | 0.843 | 0.135 | 0.012 | 0.005 | 0.005 | 0.157 |
| -87 | 0.842 | 0.136 | 0.012 | 0.005 | 0.005 | 0.158 |
| -86 | 0.842 | 0.136 | 0.013 | 0.005 | 0.005 | 0.158 |
| -85 | 0.841 | 0.137 | 0.013 | 0.005 | 0.005 | 0.159 |
| -84 | 0.840 | 0.137 | 0.013 | 0.005 | 0.005 | 0.160 |
| -83 | 0.840 | 0.138 | 0.013 | 0.005 | 0.005 | 0.160 |
| -82 | 0.839 | 0.138 | 0.013 | 0.005 | 0.005 | 0.161 |
| -81 | 0.839 | 0.139 | 0.013 | 0.005 | 0.005 | 0.161 |
| -80 | 0.838 | 0.139 | 0.013 | 0.005 | 0.005 | 0.162 |
| -79 | 0.838 | 0.140 | 0.013 | 0.005 | 0.005 | 0.162 |
| -78 | 0.837 | 0.140 | 0.013 | 0.005 | 0.005 | 0.163 |
| -77 | 0.837 | 0.141 | 0.013 | 0.005 | 0.005 | 0.163 |
| -76 | 0.836 | 0.141 | 0.013 | 0.005 | 0.005 | 0.164 |
| -75 | 0.835 | 0.142 | 0.013 | 0.005 | 0.005 | 0.165 |
| -74 | 0.835 | 0.142 | 0.013 | 0.005 | 0.005 | 0.165 |
| -73 | 0.835 | 0.142 | 0.013 | 0.005 | 0.005 | 0.165 |
| -72 | 0.835 | 0.142 | 0.013 | 0.005 | 0.005 | 0.165 |
| -71 | 0.836 | 0.142 | 0.013 | 0.005 | 0.005 | 0.164 |
| -70 | 0.836 | 0.141 | 0.013 | 0.005 | 0.005 | 0.164 |
| -69 | 0.837 | 0.141 | 0.013 | 0.005 | 0.005 | 0.163 |
| -68 | 0.836 | 0.141 | 0.013 | 0.005 | 0.005 | 0.164 |
| -67 | 0.836 | 0.141 | 0.013 | 0.005 | 0.005 | 0.164 |
| -66 | 0.835 | 0.142 | 0.013 | 0.005 | 0.005 | 0.165 |
| -65 | 0.834 | 0.143 | 0.013 | 0.005 | 0.005 | 0.166 |
| -64 | 0.833 | 0.144 | 0.013 | 0.005 | 0.005 | 0.167 |
| -63 | 0.832 | 0.145 | 0.013 | 0.005 | 0.005 | 0.168 |
| -62 | 0.831 | 0.146 | 0.014 | 0.005 | 0.005 | 0.169 |
| -61 | 0.829 | 0.147 | 0.014 | 0.005 | 0.005 | 0.171 |
| -60 | 0.828 | 0.147 | 0.014 | 0.005 | 0.005 | 0.172 |
| -59 | 0.828 | 0.148 | 0.014 | 0.005 | 0.005 | 0.172 |
| -58 | 0.827 | 0.149 | 0.014 | 0.005 | 0.005 | 0.173 |
| -57 | 0.826 | 0.149 | 0.014 | 0.005 | 0.005 | 0.174 |
| -56 | 0.826 | 0.150 | 0.014 | 0.005 | 0.005 | 0.174 |
| -55 | 0.826 | 0.150 | 0.014 | 0.005 | 0.005 | 0.174 |
| -54 | 0.826 | 0.150 | 0.014 | 0.005 | 0.005 | 0.174 |
| -53 | 0.826 | 0.150 | 0.014 | 0.005 | 0.005 | 0.174 |
| -52 | 0.826 | 0.150 | 0.014 | 0.005 | 0.005 | 0.174 |
| -51 | 0.826 | 0.150 | 0.014 | 0.005 | 0.005 | 0.174 |
| -50 | 0.826 | 0.150 | 0.014 | 0.005 | 0.005 | 0.174 |
| -49 | 0.826 | 0.150 | 0.014 | 0.005 | 0.005 | 0.174 |
| -48 | 0.826 | 0.150 | 0.014 | 0.005 | 0.005 | 0.174 |
| -47 | 0.825 | 0.150 | 0.014 | 0.005 | 0.005 | 0.175 |
| -46 | 0.825 | 0.150 | 0.014 | 0.005 | 0.005 | 0.175 |
| -45 | 0.825 | 0.150 | 0.014 | 0.005 | 0.005 | 0.175 |
| -44 | 0.825 | 0.150 | 0.014 | 0.005 | 0.005 | 0.175 |
| -43 | 0.825 | 0.151 | 0.014 | 0.005 | 0.005 | 0.175 |
| -42 | 0.824 | 0.151 | 0.014 | 0.005 | 0.006 | 0.176 |
| -41 | 0.824 | 0.151 | 0.014 | 0.005 | 0.006 | 0.176 |
| -40 | 0.824 | 0.151 | 0.014 | 0.005 | 0.006 | 0.176 |
| -39 | 0.824 | 0.151 | 0.014 | 0.005 | 0.006 | 0.176 |
| -38 | 0.824 | 0.151 | 0.014 | 0.005 | 0.006 | 0.176 |
| -37 | 0.825 | 0.151 | 0.014 | 0.005 | 0.005 | 0.175 |
| -36 | 0.825 | 0.151 | 0.014 | 0.005 | 0.005 | 0.175 |
| -35 | 0.825 | 0.151 | 0.014 | 0.005 | 0.005 | 0.175 |
| -34 | 0.824 | 0.151 | 0.014 | 0.005 | 0.006 | 0.176 |
| -33 | 0.823 | 0.152 | 0.014 | 0.005 | 0.006 | 0.177 |
| -32 | 0.822 | 0.153 | 0.014 | 0.005 | 0.006 | 0.178 |
| -31 | 0.821 | 0.154 | 0.014 | 0.005 | 0.006 | 0.179 |
| -30 | 0.819 | 0.155 | 0.015 | 0.005 | 0.006 | 0.181 |
| -29 | 0.819 | 0.156 | 0.015 | 0.005 | 0.006 | 0.181 |
| -28 | 0.818 | 0.156 | 0.015 | 0.005 | 0.006 | 0.182 |
| -27 | 0.818 | 0.156 | 0.015 | 0.005 | 0.006 | 0.182 |
| -26 | 0.818 | 0.156 | 0.015 | 0.005 | 0.006 | 0.182 |
| -25 | 0.818 | 0.156 | 0.015 | 0.005 | 0.006 | 0.182 |
| -24 | 0.818 | 0.156 | 0.015 | 0.005 | 0.006 | 0.182 |
| -23 | 0.818 | 0.156 | 0.015 | 0.005 | 0.006 | 0.182 |
| -22 | 0.818 | 0.156 | 0.015 | 0.005 | 0.006 | 0.182 |
| -21 | 0.818 | 0.156 | 0.015 | 0.005 | 0.006 | 0.182 |
| -20 | 0.817 | 0.157 | 0.015 | 0.005 | 0.006 | 0.183 |
| -19 | 0.817 | 0.157 | 0.015 | 0.005 | 0.006 | 0.183 |
| -18 | 0.816 | 0.158 | 0.015 | 0.005 | 0.006 | 0.184 |
| -17 | 0.815 | 0.159 | 0.015 | 0.005 | 0.006 | 0.185 |
| -16 | 0.814 | 0.160 | 0.015 | 0.006 | 0.006 | 0.186 |
| -15 | 0.812 | 0.161 | 0.015 | 0.006 | 0.006 | 0.188 |
| -14 | 0.811 | 0.162 | 0.015 | 0.006 | 0.006 | 0.189 |
| -13 | 0.810 | 0.163 | 0.016 | 0.006 | 0.006 | 0.190 |
| -12 | 0.809 | 0.164 | 0.016 | 0.006 | 0.006 | 0.191 |
| -11 | 0.809 | 0.164 | 0.016 | 0.006 | 0.006 | 0.191 |
| -10 | 0.809 | 0.164 | 0.016 | 0.006 | 0.006 | 0.191 |
| -9 | 0.810 | 0.163 | 0.016 | 0.006 | 0.006 | 0.190 |
| -8 | 0.811 | 0.162 | 0.015 | 0.006 | 0.006 | 0.189 |
| -7 | 0.812 | 0.161 | 0.015 | 0.006 | 0.006 | 0.188 |
| -6 | 0.812 | 0.161 | 0.015 | 0.006 | 0.006 | 0.188 |
| -5 | 0.811 | 0.162 | 0.015 | 0.006 | 0.006 | 0.189 |
| -4 | 0.809 | 0.164 | 0.016 | 0.006 | 0.006 | 0.191 |
| -3 | 0.805 | 0.167 | 0.016 | 0.006 | 0.006 | 0.195 |
| -2 | 0.798 | 0.173 | 0.017 | 0.006 | 0.007 | 0.202 |
| -1 | 0.788 | 0.181 | 0.018 | 0.006 | 0.007 | 0.212 |
| 0 | 0.778 | 0.189 | 0.019 | 0.007 | 0.007 | 0.222 |
| 1 | 0.768 | 0.197 | 0.020 | 0.007 | 0.008 | 0.232 |
| 2 | 0.761 | 0.203 | 0.020 | 0.008 | 0.008 | 0.239 |
| 3 | 0.758 | 0.206 | 0.021 | 0.008 | 0.008 | 0.242 |
| 4 | 0.759 | 0.205 | 0.021 | 0.008 | 0.008 | 0.241 |
| 5 | 0.763 | 0.201 | 0.020 | 0.007 | 0.008 | 0.237 |
| 6 | 0.769 | 0.196 | 0.020 | 0.007 | 0.008 | 0.231 |
| 7 | 0.776 | 0.190 | 0.019 | 0.007 | 0.007 | 0.224 |
| 8 | 0.783 | 0.185 | 0.018 | 0.007 | 0.007 | 0.217 |
| 9 | 0.789 | 0.180 | 0.018 | 0.006 | 0.007 | 0.211 |
| 10 | 0.793 | 0.177 | 0.017 | 0.006 | 0.007 | 0.207 |
| 11 | 0.795 | 0.175 | 0.017 | 0.006 | 0.007 | 0.205 |
| 12 | 0.796 | 0.174 | 0.017 | 0.006 | 0.007 | 0.204 |
| 13 | 0.796 | 0.174 | 0.017 | 0.006 | 0.007 | 0.204 |
| 14 | 0.796 | 0.174 | 0.017 | 0.006 | 0.007 | 0.204 |
| 15 | 0.796 | 0.175 | 0.017 | 0.006 | 0.007 | 0.204 |
| 16 | 0.796 | 0.174 | 0.017 | 0.006 | 0.007 | 0.204 |
| 17 | 0.797 | 0.174 | 0.017 | 0.006 | 0.007 | 0.203 |
| 18 | 0.797 | 0.173 | 0.017 | 0.006 | 0.007 | 0.203 |
| 19 | 0.798 | 0.173 | 0.017 | 0.006 | 0.007 | 0.202 |
| 20 | 0.798 | 0.173 | 0.017 | 0.006 | 0.007 | 0.202 |
| 21 | 0.797 | 0.173 | 0.017 | 0.006 | 0.007 | 0.203 |
| 22 | 0.797 | 0.173 | 0.017 | 0.006 | 0.007 | 0.203 |
| 23 | 0.797 | 0.174 | 0.017 | 0.006 | 0.007 | 0.203 |
| 24 | 0.796 | 0.174 | 0.017 | 0.006 | 0.007 | 0.204 |
| 25 | 0.796 | 0.175 | 0.017 | 0.006 | 0.007 | 0.204 |
| 26 | 0.795 | 0.175 | 0.017 | 0.006 | 0.007 | 0.205 |
| 27 | 0.794 | 0.176 | 0.017 | 0.006 | 0.007 | 0.206 |
| 28 | 0.792 | 0.178 | 0.017 | 0.006 | 0.007 | 0.208 |
| 29 | 0.791 | 0.179 | 0.017 | 0.006 | 0.007 | 0.209 |
| 30 | 0.789 | 0.180 | 0.018 | 0.006 | 0.007 | 0.211 |
| 31 | 0.789 | 0.180 | 0.018 | 0.006 | 0.007 | 0.211 |
| 32 | 0.789 | 0.181 | 0.018 | 0.006 | 0.007 | 0.211 |
| 33 | 0.789 | 0.180 | 0.018 | 0.006 | 0.007 | 0.211 |
| 34 | 0.790 | 0.179 | 0.018 | 0.006 | 0.007 | 0.210 |
| 35 | 0.790 | 0.179 | 0.017 | 0.006 | 0.007 | 0.210 |
| 36 | 0.791 | 0.179 | 0.017 | 0.006 | 0.007 | 0.209 |
| 37 | 0.791 | 0.179 | 0.017 | 0.006 | 0.007 | 0.209 |
| 38 | 0.790 | 0.179 | 0.017 | 0.006 | 0.007 | 0.210 |
| 39 | 0.789 | 0.180 | 0.018 | 0.006 | 0.007 | 0.211 |
| 40 | 0.789 | 0.180 | 0.018 | 0.006 | 0.007 | 0.211 |
| 41 | 0.788 | 0.181 | 0.018 | 0.006 | 0.007 | 0.212 |
| 42 | 0.787 | 0.182 | 0.018 | 0.006 | 0.007 | 0.213 |
| 43 | 0.787 | 0.182 | 0.018 | 0.007 | 0.007 | 0.213 |
| 44 | 0.787 | 0.182 | 0.018 | 0.007 | 0.007 | 0.213 |
| 45 | 0.787 | 0.182 | 0.018 | 0.007 | 0.007 | 0.213 |
| 46 | 0.788 | 0.181 | 0.018 | 0.006 | 0.007 | 0.212 |
| 47 | 0.788 | 0.181 | 0.018 | 0.006 | 0.007 | 0.212 |
| 48 | 0.789 | 0.180 | 0.018 | 0.006 | 0.007 | 0.211 |
| 49 | 0.789 | 0.180 | 0.018 | 0.006 | 0.007 | 0.211 |
| 50 | 0.789 | 0.180 | 0.018 | 0.006 | 0.007 | 0.211 |
| 51 | 0.789 | 0.180 | 0.018 | 0.006 | 0.007 | 0.211 |
| 52 | 0.789 | 0.180 | 0.018 | 0.006 | 0.007 | 0.211 |
| 53 | 0.790 | 0.179 | 0.018 | 0.006 | 0.007 | 0.210 |
| 54 | 0.790 | 0.179 | 0.017 | 0.006 | 0.007 | 0.210 |
| 55 | 0.790 | 0.180 | 0.018 | 0.006 | 0.007 | 0.210 |
| 56 | 0.789 | 0.180 | 0.018 | 0.006 | 0.007 | 0.211 |
| 57 | 0.787 | 0.182 | 0.018 | 0.006 | 0.007 | 0.213 |
| 58 | 0.786 | 0.183 | 0.018 | 0.007 | 0.007 | 0.214 |
| 59 | 0.785 | 0.184 | 0.018 | 0.007 | 0.007 | 0.215 |
| 60 | 0.784 | 0.184 | 0.018 | 0.007 | 0.007 | 0.216 |
| 61 | 0.785 | 0.184 | 0.018 | 0.007 | 0.007 | 0.215 |
| 62 | 0.786 | 0.183 | 0.018 | 0.007 | 0.007 | 0.214 |
| 63 | 0.786 | 0.182 | 0.018 | 0.007 | 0.007 | 0.214 |
| 64 | 0.787 | 0.182 | 0.018 | 0.007 | 0.007 | 0.213 |
| 65 | 0.786 | 0.182 | 0.018 | 0.007 | 0.007 | 0.214 |
| 66 | 0.786 | 0.183 | 0.018 | 0.007 | 0.007 | 0.214 |
| 67 | 0.785 | 0.184 | 0.018 | 0.007 | 0.007 | 0.215 |
| 68 | 0.784 | 0.184 | 0.018 | 0.007 | 0.007 | 0.216 |
| 69 | 0.783 | 0.185 | 0.018 | 0.007 | 0.007 | 0.217 |
| 70 | 0.783 | 0.185 | 0.018 | 0.007 | 0.007 | 0.217 |
| 71 | 0.783 | 0.185 | 0.018 | 0.007 | 0.007 | 0.217 |
| 72 | 0.784 | 0.185 | 0.018 | 0.007 | 0.007 | 0.216 |
| 73 | 0.784 | 0.184 | 0.018 | 0.007 | 0.007 | 0.216 |
| 74 | 0.784 | 0.184 | 0.018 | 0.007 | 0.007 | 0.216 |
| 75 | 0.784 | 0.184 | 0.018 | 0.007 | 0.007 | 0.216 |
| 76 | 0.783 | 0.185 | 0.018 | 0.007 | 0.007 | 0.217 |
| 77 | 0.782 | 0.186 | 0.018 | 0.007 | 0.007 | 0.218 |
| 78 | 0.780 | 0.187 | 0.018 | 0.007 | 0.007 | 0.220 |
| 79 | 0.779 | 0.188 | 0.019 | 0.007 | 0.007 | 0.221 |
| 80 | 0.778 | 0.189 | 0.019 | 0.007 | 0.007 | 0.222 |
| 81 | 0.778 | 0.189 | 0.019 | 0.007 | 0.007 | 0.222 |
| 82 | 0.778 | 0.189 | 0.019 | 0.007 | 0.007 | 0.222 |
| 83 | 0.777 | 0.190 | 0.019 | 0.007 | 0.007 | 0.223 |
| 84 | 0.777 | 0.190 | 0.019 | 0.007 | 0.007 | 0.223 |
| 85 | 0.776 | 0.191 | 0.019 | 0.007 | 0.007 | 0.224 |
| 86 | 0.776 | 0.191 | 0.019 | 0.007 | 0.007 | 0.224 |
| 87 | 0.776 | 0.191 | 0.019 | 0.007 | 0.007 | 0.224 |
| 88 | 0.776 | 0.191 | 0.019 | 0.007 | 0.007 | 0.224 |
| 89 | 0.777 | 0.190 | 0.019 | 0.007 | 0.007 | 0.223 |
| 90 | 0.777 | 0.190 | 0.019 | 0.007 | 0.007 | 0.223 |
| 91 | 0.778 | 0.189 | 0.019 | 0.007 | 0.007 | 0.222 |
| 92 | 0.778 | 0.189 | 0.019 | 0.007 | 0.007 | 0.222 |
| 93 | 0.778 | 0.189 | 0.019 | 0.007 | 0.007 | 0.222 |
| 94 | 0.778 | 0.189 | 0.019 | 0.007 | 0.007 | 0.222 |
| 95 | 0.778 | 0.189 | 0.019 | 0.007 | 0.007 | 0.222 |
| 96 | 0.778 | 0.189 | 0.019 | 0.007 | 0.007 | 0.222 |
| 97 | 0.778 | 0.189 | 0.019 | 0.007 | 0.007 | 0.222 |
| 98 | 0.778 | 0.189 | 0.019 | 0.007 | 0.007 | 0.222 |
| 99 | 0.778 | 0.189 | 0.019 | 0.007 | 0.007 | 0.222 |
| 100 | 0.778 | 0.189 | 0.019 | 0.007 | 0.007 | 0.222 |
| 101 | 0.777 | 0.190 | 0.019 | 0.007 | 0.007 | 0.223 |
| 102 | 0.777 | 0.190 | 0.019 | 0.007 | 0.007 | 0.223 |
| 103 | 0.776 | 0.191 | 0.019 | 0.007 | 0.007 | 0.224 |
| 104 | 0.775 | 0.191 | 0.019 | 0.007 | 0.007 | 0.225 |
| 105 | 0.775 | 0.192 | 0.019 | 0.007 | 0.007 | 0.225 |
| 106 | 0.774 | 0.192 | 0.019 | 0.007 | 0.008 | 0.226 |
| 107 | 0.774 | 0.192 | 0.019 | 0.007 | 0.008 | 0.226 |
| 108 | 0.774 | 0.192 | 0.019 | 0.007 | 0.008 | 0.226 |
| 109 | 0.774 | 0.192 | 0.019 | 0.007 | 0.008 | 0.226 |
| 110 | 0.774 | 0.192 | 0.019 | 0.007 | 0.008 | 0.226 |
| 111 | 0.774 | 0.192 | 0.019 | 0.007 | 0.008 | 0.226 |
| 112 | 0.774 | 0.192 | 0.019 | 0.007 | 0.008 | 0.226 |
| 113 | 0.774 | 0.192 | 0.019 | 0.007 | 0.008 | 0.226 |
| 114 | 0.773 | 0.193 | 0.019 | 0.007 | 0.008 | 0.227 |
| 115 | 0.772 | 0.194 | 0.019 | 0.007 | 0.008 | 0.228 |
| 116 | 0.771 | 0.195 | 0.019 | 0.007 | 0.008 | 0.229 |
| 117 | 0.770 | 0.196 | 0.020 | 0.007 | 0.008 | 0.230 |
| 118 | 0.770 | 0.196 | 0.020 | 0.007 | 0.008 | 0.230 |
| 119 | 0.771 | 0.195 | 0.019 | 0.007 | 0.008 | 0.229 |
| 120 | 0.772 | 0.194 | 0.019 | 0.007 | 0.008 | 0.228 |

Table A4: Table of modelled categorical responses for physical contacts with 18 to 69 year olds and its translation to a probability of having had some contacts, i.e. summing response categories 2 to 5.

Appendix 4: testing behavioural differences between ChAdOx1 versus the BNT162b2 vaccines

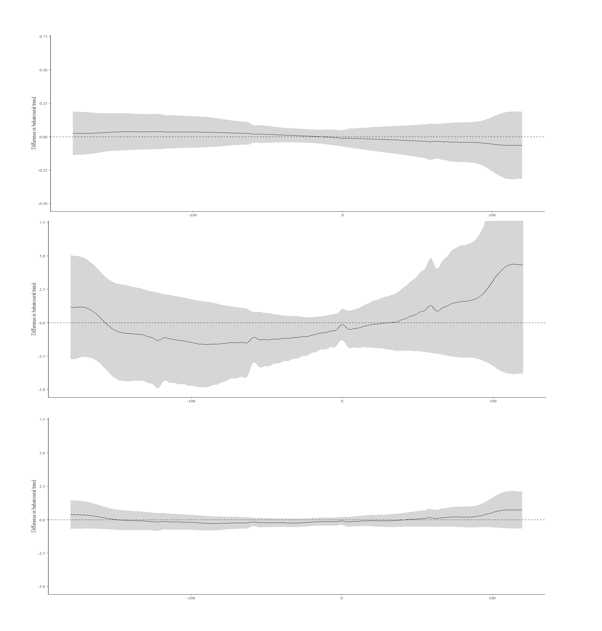

Table A5: differences in smooths between ChAdOx1 versus the BNT162b2 vaccines for adults aged 18-64 years. Top panel: others in own home in the past 7 days. Middle panel: physical contacts with 18 to 69 year-olds in the past 7 days. Bottom panel: Socially-distanced contacts with 18 to 69 year-olds in the past 7 days. No evidence to suggest behavioural trends diverge between individuals that received either ChAdOx1 versus the BNT162b2 vaccines.

Appendix 5: behavioural response to second vaccination

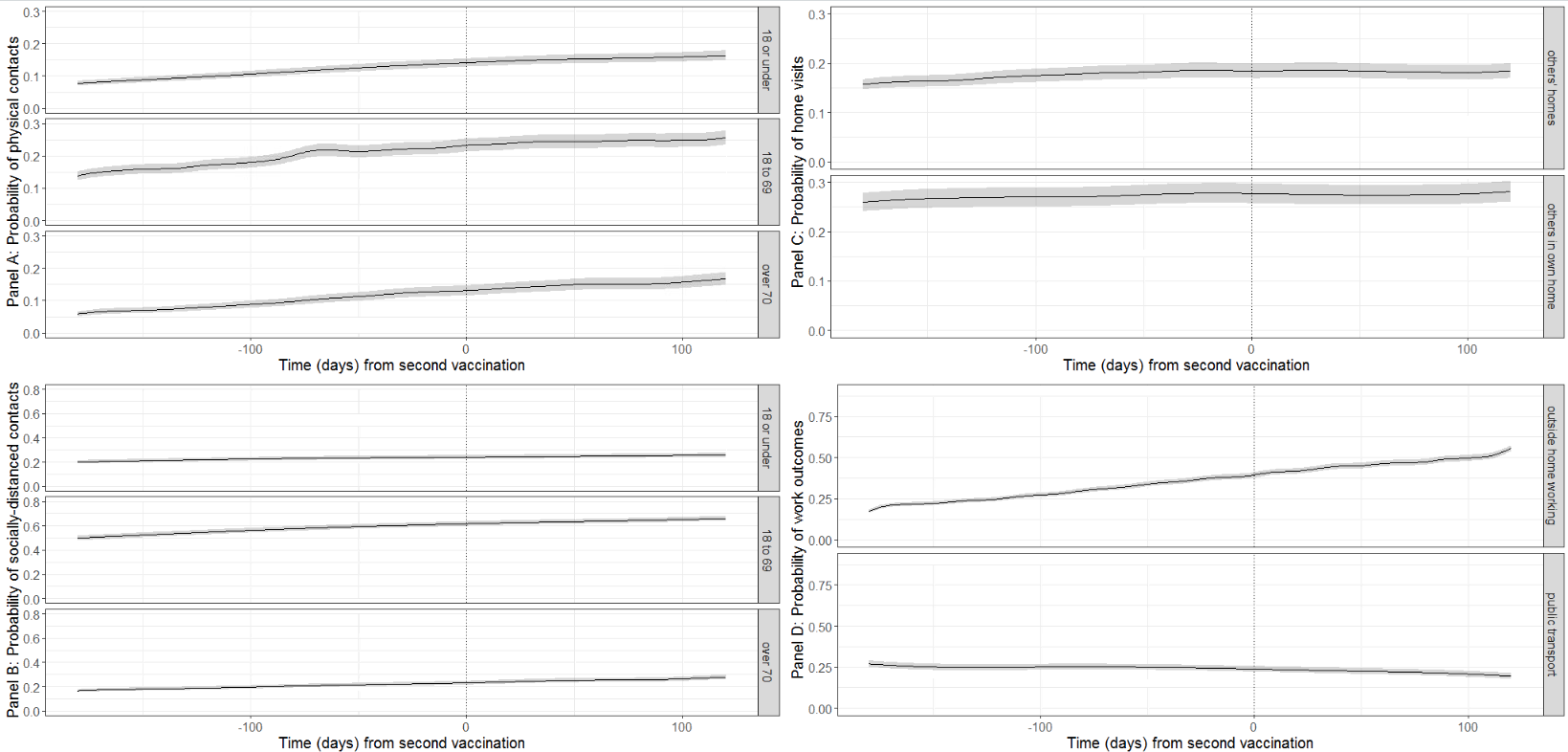

Figure A6: Probabilities of behavioural outcomes for individuals aged 18-64y by time from first vaccination, second dose. Top left (panel A): past 7-day reported physical, outside of household contacts; bottom left (Panel B): past 7-day reported socially-distanced, outside of household contacts; top right (Panel C): past 7-day reported home visits; bottom right (Panel D): past 7-day reported work outcomes for those that are working or in education. Dotted line shows day of own first vaccination. “18 or under”, “18 to 69” and “over 70” denote the ages of the people that individuals in the sample had contact with.

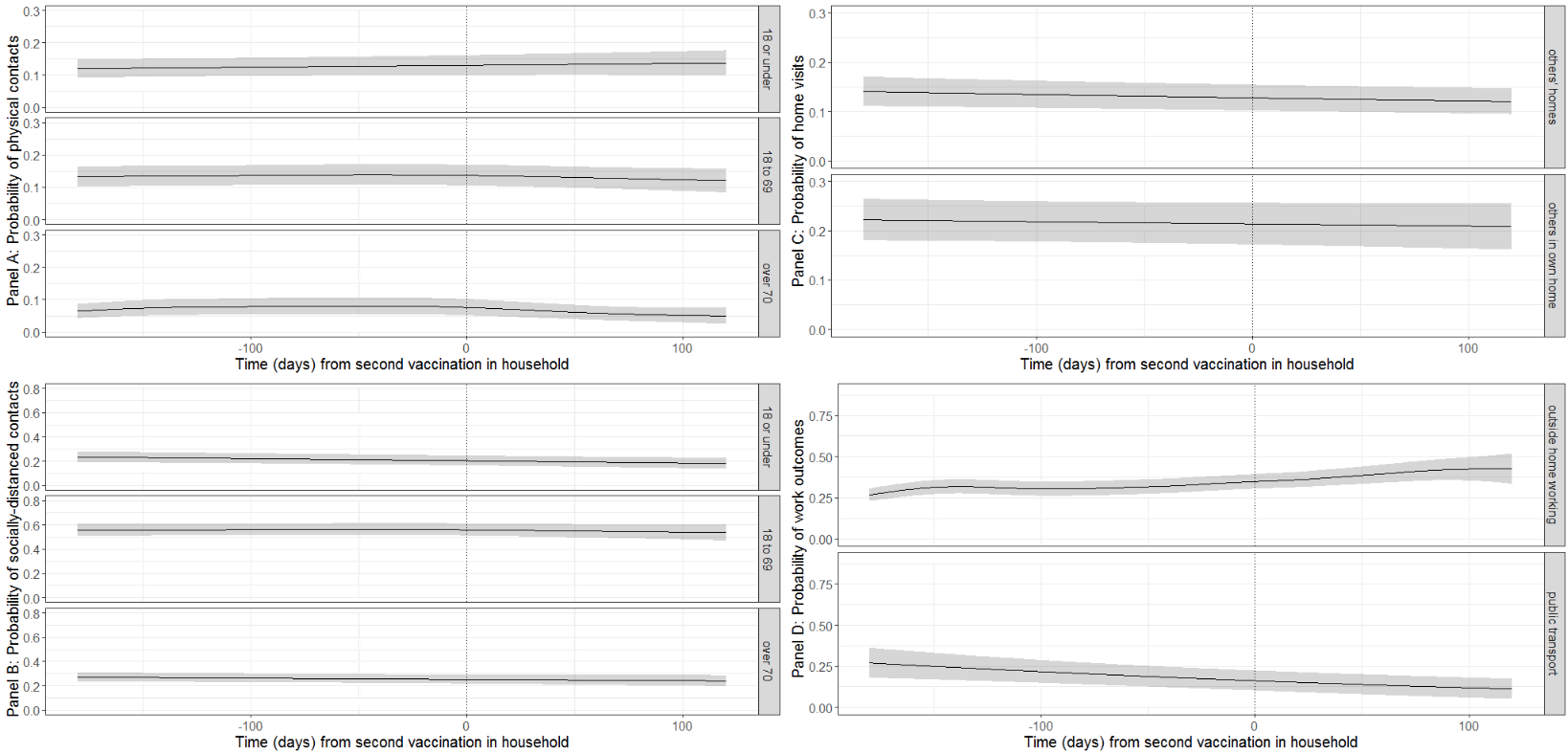

Figure A7: Probabilities of behavioural outcomes for unvaccinated individuals aged 18-64y by time to vaccination of the first vulnerable person in the household, second dose. Top left (panel A): past 7-day reported physical, outside of household contacts; bottom left (Panel B): past 7-day reported socially-distanced, outside of household contacts; top right (Panel C): past 7-day reported home visits; bottom right (Panel D): past 7-day reported work outcomes for those that are working or in education. Dotted line shows day of own first vaccination. “18 or under”, “18 to 69” and “over 70” denote the ages of the people that individuals in the sample had contact with.

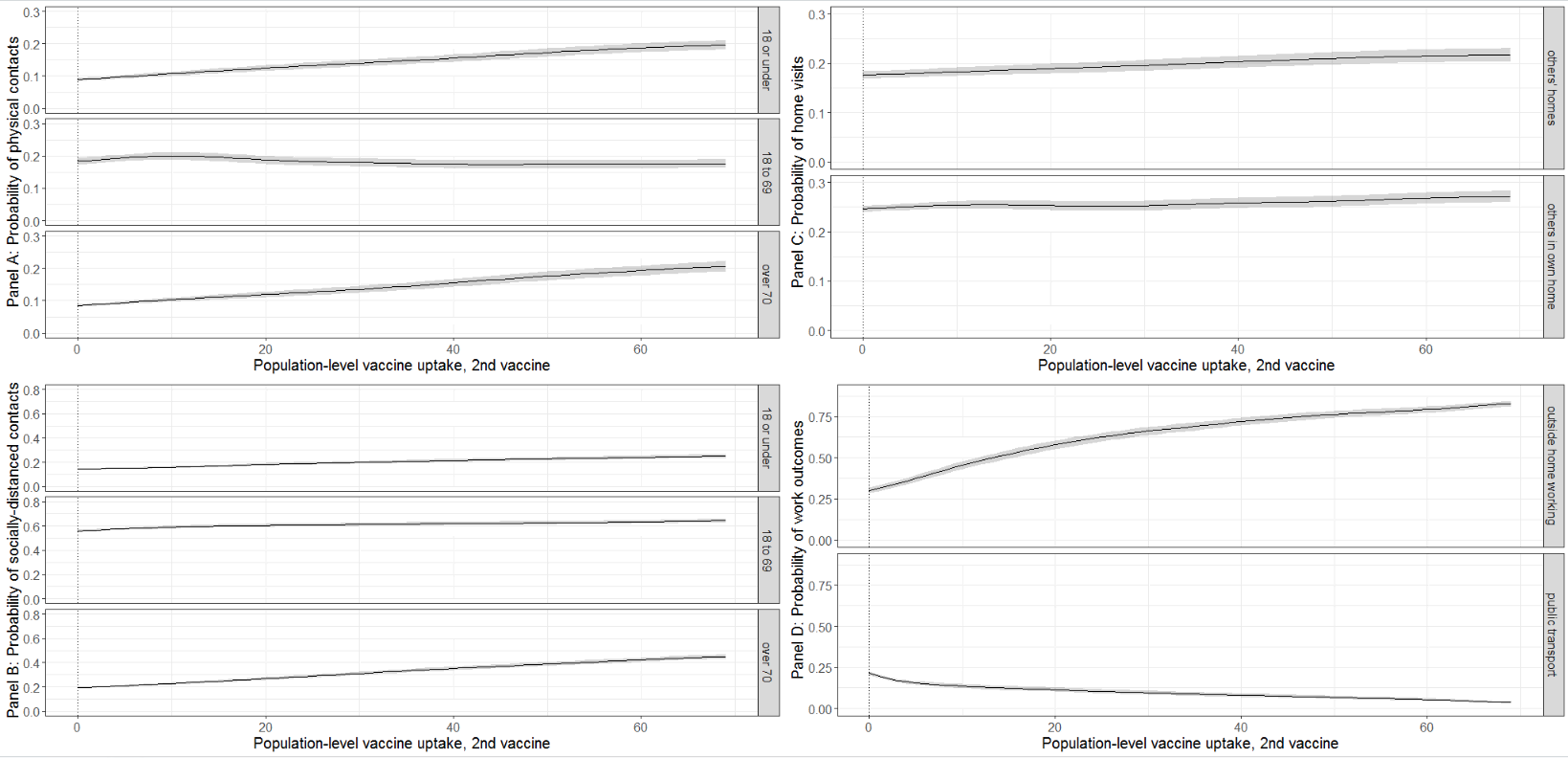

Figure A8: Probabilities of behavioural outcomes by population level vaccination %, second dose. Top left (panel A): past 7-day reported physical, outside of household contacts; bottom left (Panel B): past 7-day reported socially-distanced, outside of household contacts; top right (Panel C): past 7-day reported home visits; bottom right (Panel D): past 7-day reported work outcomes for those that are working or in education. “18 or under”, “18 to 69” and “over 70” denote the ages of the people that individuals in the sample had contact with.

Appendix 6: Analysis of population level vaccination restricted to the sample used for response to own vaccination

Fig A9 shows the variation in 10 behavioural outcomes as a function of the rate of first vaccination in the population, using the sample of vaccinated individuals as per individual vaccination.

The probability of outside of the household - physical and socially-distanced - contacts increased as population level vaccination increased (Fig A9 panels A and B). In contrast to the full sample, an initial peak in the probability of contacts in the first 25% of population vaccination is not observed. Probabilities of home visits appeared to be stable as population vaccination rates increased (Fig A9 panel C). Probabilities of both working at home and taking public transport decreased as population vaccination rates increased (Fig A9 panel D).

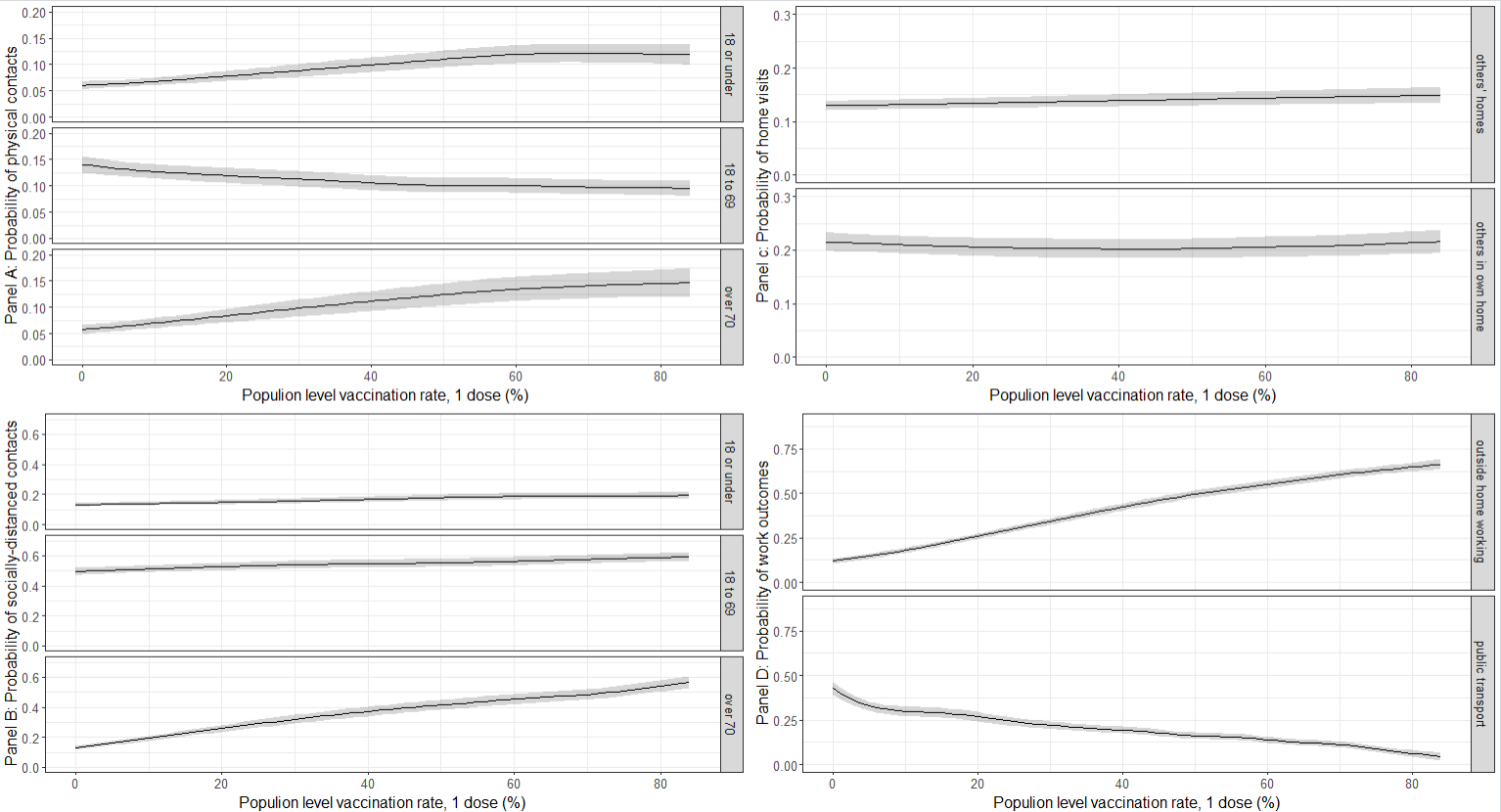

Figure A9: Probabilities of behavioural outcomes by population level vaccination %, first dose. Top left (panel A): past 7-day reported physical, outside of household contacts; bottom left (Panel B): past 7-day reported socially-distanced, outside of household contacts; top right (Panel C): past 7-day reported home visits; bottom right (Panel D): past 7-day reported work outcomes for those that are working or in education. “18 or under”, “18 to 69” and “over 70” denote the ages of the people that individuals in the sample had contact with.

Appendix 7: Calendar time by region/country (9 regions in England and Northern Ireland, Scotland, and Wales) for physical contacts with under 18 year-olds; vaccinated individuals

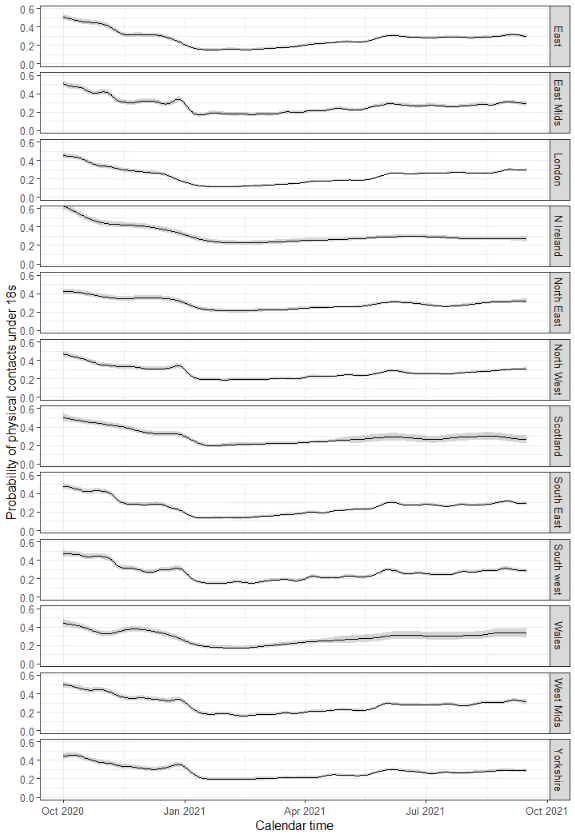

Appendix 8: counts of responses to outcomes, own vaccination of 18 to 64 year-olds

| Outcome | Behaviour captured by binary variable | Counts | | | | | |
| --- | --- | --- | --- | --- | --- | --- | --- |
|  |  | zero | any | n | missing | denominator | % any |
| Physical contacts with under 18s | Any  past 7-day contacts outside the household | 1418281 | 421630 | 1839911 | 43306 | 1796605 | 23.5% |
| Physical contacts with 18 to 69 years | Any past 7-day contacts outside the household | 1153840 | 686071 | 1839911 | 41423 | 1798488 | 38.1% |
| Physical contacts with over 70s | Any past 7-day contacts outside the household | 1524370 | 315541 | 1839911 | 45096 | 1794815 | 17.6% |
| Socially-distanced contacts with under 18s | Any past 7-day contacts outside the household | 1226960 | 612951 | 1839911 | 46395 | 1793516 | 34.2% |
| Socially-distanced contacts with 18 to 69 years | Any past 7-day contacts outside the household | 483189 | 1356722 | 1839911 | 41775 | 1798136 | 75.5% |
| Socially-distanced contacts with over 70s | Any past 7-day contacts outside the household | 1226525 | 613386 | 1839911 | 46568 | 1793343 | 34.2% |
| Visits to others' homes | Any past 7-day home visits | 1226558 | 613353 | 1839911 | 156288 | 1683623 | 36.4% |
| Others' visits to own home | Any past 7-day home visits | 1153551 | 686360 | 1839911 | 157983 | 1681928 | 40.8% |
| Working/studying at home | Any past 7-day working at home | 553886 | 455669 | 1009555 | 32215 | 977340 | 46.6% |
| Taking public transport to work/place of education | Any past 7-day public transport | 725771 | 76021 | 1009555 | 207762 | 801793 | 9.5% |

Counts of self-reported behaviours across all outcomes. The numerator is the number of observations for which “any” was reported and the denominator is the total number of observations in that analysis. Denominators for work variables restricted to those reporting working or in education. Zero – counts of “none”, any – counts of “any”, n – sample size in analysis, missing – missing outcome (not reported), denominator – sample size less missing observations, % any – percentage of observations reporting “any”.

Appendix 9: counts of responses to outcomes, household vaccination of 18 to 64 year-olds

| Outcome | Behaviour captured by binary variable | Counts | | | | | |
| --- | --- | --- | --- | --- | --- | --- | --- |
|  |  | zero | any | n | missing | denominator | % any |
| Physical contacts with under 18s | Any  past 7-day contacts outside the household | 42198 | 11648 | 53846 | 1900 | 51946 | 22.4% |
| Physical contacts with 18 to 69 years | Any past 7-day contacts outside the household | 36745 | 17101 | 53846 | 1805 | 52041 | 32.9% |
| Physical contacts with over 70s | Any past 7-day contacts outside the household | 46099 | 7747 | 53846 | 1957 | 51889 | 14.9% |
| Socially-distanced contacts with under 18s | Any past 7-day contacts outside the household | 38121 | 15725 | 53846 | 2011 | 51835 | 30.3% |
| Socially-distanced contacts with 18 to 69 years | Any past 7-day contacts outside the household | 16334 | 37512 | 53846 | 1787 | 52059 | 72.1% |
| Socially-distanced contacts with over 70s | Any past 7-day contacts outside the household | 36947 | 16899 | 53846 | 2042 | 51804 | 32.6% |
| Visits to others' homes | Any past 7-day home visits | 37235 | 16611 | 53846 | 7261 | 46585 | 35.7% |
| Others' visits to own home | Any past 7-day home visits | 34912 | 18934 | 53846 | 7329 | 46517 | 40.7% |
| Working/studying at home | Any past 7-day working at home | 37810 | 29721 | 67531 | 3138 | 64393 | 46.2% |
| Taking public transport to work/place of education | Any past 7-day public transport | 63898 | 3633 | 67531 | 25925 | 41606 | 8.7% |

Counts of self-reported behaviours across all outcomes. The numerator is the number of observations for which “any” was reported and the denominator is the total number of observations in that analysis. Denominators for work variables restricted to those reporting working or in education. Zero – counts of “none”, any – counts of “any”, n – sample size in analysis, missing – missing outcome (not reported), denominator – sample size less missing observations, % any – percentage of observations reporting “any”.

Appendix 10: counts of responses to outcomes, population level vaccination of all ages

| Outcome | Behaviour captured by binary variable | Counts | | | | | |
| --- | --- | --- | --- | --- | --- | --- | --- |
|  |  | zero | any | n | missing | denominator | % any |
| Physical contacts with under 18s | Any  past 7-day contacts outside the household | 3200446 | 1308309 | 4508755 | 99288 | 4409467 | 29.7% |
| Physical contacts with 18 to 69 years | Any past 7-day contacts outside the household | 2755714 | 1753041 | 4508755 | 95977 | 4412778 | 39.7% |
| Physical contacts with over 70s | Any past 7-day contacts outside the household | 3700848 | 807907 | 4508755 | 103869 | 4404886 | 18.3% |
| Socially-distanced contacts with under 18s | Any past 7-day contacts outside the household | 2913325 | 1595430 | 4508755 | 106461 | 4402294 | 36.2% |
| Socially-distanced contacts with 18 to 69 years | Any past 7-day contacts outside the household | 1324056 | 3184699 | 4508755 | 97360 | 4411395 | 72.2% |
| Socially-distanced contacts with over 70s | Any past 7-day contacts outside the household | 2994133 | 1514622 | 4508755 | 107406 | 4401349 | 34.4% |
| Visits to others' homes | Any past 7-day home visits | 3057637 | 1451118 | 4508755 | 351187 | 4157568 | 34.9% |
| Others' visits to own home | Any past 7-day home visits | 2826878 | 1681877 | 4508755 | 354991 | 4153764 | 40.5% |
| Working/studying at home | Any past 7-day working at home | 1819799 | 885522 | 2705321 | 656717 | 2048604 | 43.2% |
| Taking public transport to work/place of education | Any past 7-day public transport | 2509774 | 195547 | 2705321 | 934895 | 1770426 | 11.0% |

Counts of self-reported behaviours across all outcomes. The numerator is the number of observations for which “any” was reported and the denominator is the total number of observations in that analysis. Denominators for work variables restricted to those reporting working or in education. Zero – counts of “none”, any – counts of “any”, n – sample size in analysis, missing – missing outcome (not reported), denominator – sample size less missing observations, % any – percentage of observations reporting “any”.
